# The *Soifua Manuia* reference panel with 2,570 Samoan haplotypes improves genotype imputation quality among Samoans

**DOI:** 10.1101/2023.10.31.23297835

**Authors:** Jenna C. Carlson, Mohanraj Krishnan, Shuwei Liu, Kevin J. Anderson, Jerry Z. Zhang, Lauren M. Spor, Toni-Ann J. Yapp, Elizabeth A. Chiyka, Devin A. Dikec, Hong Cheng, Take Naseri, Muagututi‘a Sefuiva Reupena, Satupa‘itea Viali, Ranjan Deka, Nicola L. Hawley, Stephen T. McGarvey, Daniel E. Weeks, Ryan L. Minster

## Abstract

Genotype imputation is fundamental to association studies, and yet even gold standard panels like TOPMed are limited in the populations for which they yield good imputation. Specifically, Pacific Islanders are poorly represented in extant panels. To address this, we constructed an imputation reference panel using 1,285 Samoan individuals with whole-genome sequencing, combined with 1000 Genomes Project (1KGP) individuals, to create a reference panel that better represents Pacific Islander, specifically Samoan, genetic variation. We compared this panel to 1KGP and TOPMed-R3 panels based on imputed variants using genotyping array data for 1,834 Samoan participants who were not part of the panels. The 1KGP + 1285 Samoan panel yielded up to two times more well-imputed (r^2^ ≥ 0.80) variants than TOPMed-R3 and 1KGP and was enriched for moderate and high impact variants. There was improved imputation accuracy across the minor allele frequency (MAF) spectrum, although it was most pronounced for variants with 0.01 ≤ MAF ≤ 0.05. Imputation accuracy (r^2^) was greater for population-specific variants (high fixation index, F_ST_) and those from larger haplotypes (high LD score). However, the gain in imputation accuracy over TOPMed-R3 was largest for small haplotypes (low LD score), reflecting the Samoan panel’s ability to capture population-specific variation not well tagged by other panels. We also augmented the 1KGP reference panel with varying numbers of Samoan participants and found that panels with 24 Samoans yielded similar performance to TOPMed-R3, and panels with 48 or more Samoans included outperformed TOPMed-R3 for all variants with MAF ≥ 0.001. Meta imputation of the TOPMed-R3 and 1285 Samoan panels yielded poorer performance than the Samoan only panel. We also demonstrated that the phasing of the reference panel impacts the imputation of population-specific variants when the reference panel is composed of individuals from an isolated population and not combined with ancestrally diverse haplotypes. This study identifies variants with improved imputation using population-specific reference panels and provides a framework for constructing other population-specific reference panels.

## Introduction

Genotype imputation is fundamental to modern genome-wide association studies (GWAS), yielding a much denser association landscape in which fine-mapping of causal variants and genes can occur than in what can be obtained using genotyping array data alone. However, the utility of genotype imputation depends upon the haplotypes that make up the reference panels. This is especially critical in populations that are underrepresented in genetic research, as imputation is much poorer when the reference panel does not contain good haplotype matches to that of the target population. Existing panels like 1000 Genomes (1KGP)^1^, the Haplotype Reference Consortium (HRC)^2^, and the Trans-Omics for Precision Medicine version R3 (TOPMed-R3)^3^ panels catalog human genome variation of numerous majority populations worldwide, especially those with Western European ancestries. The TOPMed-R3 panel contains the most ancestrally diverse set of participants and has addressed disparities in representation, especially for Hispanic- and African-admixed populations, greatly improving imputation accuracy in these populations^3,4^. However, there are still populations for which these reference panels are inadequate. This is obvious in looking at the representation of Pacific Islanders, who are underrepresented in health research. In common publicly available reference panels (1KGP, HRC, and TOPMed-R3), there are no participants explicitly from Oceania, and so the totality of genetic variation present in Pacific Islanders cannot be adequately represented in genetic association studies. This may further exacerbate the health disparities faced by Pacific Islanders, who are not only underrepresented in research but have a disproportionate burden of non-communicable diseases compared to majority populations.

For example, the Pacific Island nation of Samoa, which has a unique population history consisting of founder effects and bottlenecks^5–8^, also faces some of the highest rates of non-communicable disease globally^9–13^. Despite this, most existing knowledge about disease etiology comes from majority ethnic groups, even though disease presentation, progression, and response to treatment can differ by ancestry^11,14–21^. Specific to genetic studies, Pacific Islanders more broadly are often grouped into an “other/mixed” ancestral category which itself is still <1% of GWAS participants^22^. However, because of their unique population history, there are regions of the genome for which Samoans have different variant frequencies and haplotype structure (LD) from other majority populations, making the aggregation of them with other Oceanic- or Asian-ancestry individuals inappropriate. Moreover, advances in knowledge on the genetic underpinnings of human phenotypic variation from majority populations are often assumed to apply to all populations globally, which is premature at best (but, more likely, contributing to racial health disparity)^23,24^.

To combat this, greater representation of Pacific Islanders is needed in gene-mapping studies. However, this cannot happen without additional whole-genome sequencing and/or improved genotype imputation. Evidence of this comes from studies based on cohorts of Samoans and American Samoans recruited in 1990-1995^25–27^, 2002-2003^28,29^, and 2010^12^. We have performed GWAS to dissect the genetic architecture of cardiometabolic disease traits in these cohorts.

Through this, we have identified variants of high frequency in Samoa and not appearing (at least at high frequency) in other non-Pacific-Islander populations that are associated with body mass index^30^ and lipid traits^31^. They may be at high frequency due to founding events and bottlenecks. Interestingly, these variants were not present in initial GWAS using genotyping array data only. They were discovered using imputation based on Samoan haplotypes. In the two loci harboring Samoan-specific variants^30,31^, the genetic architecture of the GWAS signal looks markedly different after imputation, with the location of the sentinel variants shifting dramatically post-imputation. The discovery of these associations offers a case study into how genetic studies in Samoans and other Pacific-Islander populations will benefit from improved imputation.

In this study, we describe the strategy we employed to create a Samoan-specific reference panel containing phased haplotypes from 1KGP participants and Samoan participants with whole-genome sequencing (WGS) from the Samoan Study, part of the TOPMed Consortium. Importantly, the Samoan Study, which consists of 1,285 participants of Samoan ancestry that are part of the TOPMed Consortium, was not included in the TOPMed-R3 reference panel. Many individuals of Samoan ancestry can also have Asian or European ancestries; to impute well in those individuals, it is necessary to include diversity such as that of the 1KGP to the panel.

We investigated this panel derived from Samoan participants with WGS and compared it to several other reference panels, to determine the extent to which adding Samoan participants to the existing 1KGP reference panel impacted imputation quality. Specifically, we sought to address these questions:

1) How does the imputation for a reference panel of 1,285 Samoans plus 1KGP compare to other existing panels?
2) How many population-specific sequences are necessary to add to 1KGP to achieve good imputation accuracy?
3) Does the phasing of the reference panel impact imputed accuracy and genotype frequency?
4) How does meta imputation of a population-specific panel with the TOPMed-R3 panel perform compared to the panels separately?

## Subjects and Methods

Participants for this study came from the *Soifua Manuia* (‘good health’) study, a population-based sample of 3,504 Samoan adults recruited in 2010^12^. Genotyping was performed for a subset of 3,182 Samoan adults using Genome-Wide Human SNP 6.0 arrays (Affymetrix) and extensive quality control was conducted via a pipeline developed by Laurie et al.^32^. Additional details about sample genotyping and genotype quality control are described in Minster et al.^30^. A subset (n=1,285) from this cohort also had WGS data from TOPMed. These 1,285 individuals were optimally selected for WGS by using the rankings from INFOSTIP^33^ to capture the cohort’s genetic diversity (http://www.cs.columbia.edu/~itsik/software/INFOSTIP.html) (Figures S1, S2). In addition to the Samoan participants, 2,504 individuals from the 1000 Genomes Project Phase III (1KGP) with WGS through TOPMed were included where appropriate.

### Construction and comparison of the reference panels

We constructed 14 in-house imputation reference panels, varying the number of Samoan participants, the inclusion of 1KGP individuals, and using different phasing strategies, to assess differences in imputation quality across a variety of settings (Figure 1, Table 1). We included high-quality (i.e., passing all QC filters and with a minimum read depth of 10), bi-allelic markers from the TOPMed freeze 9b call set.

**Figure 1.**
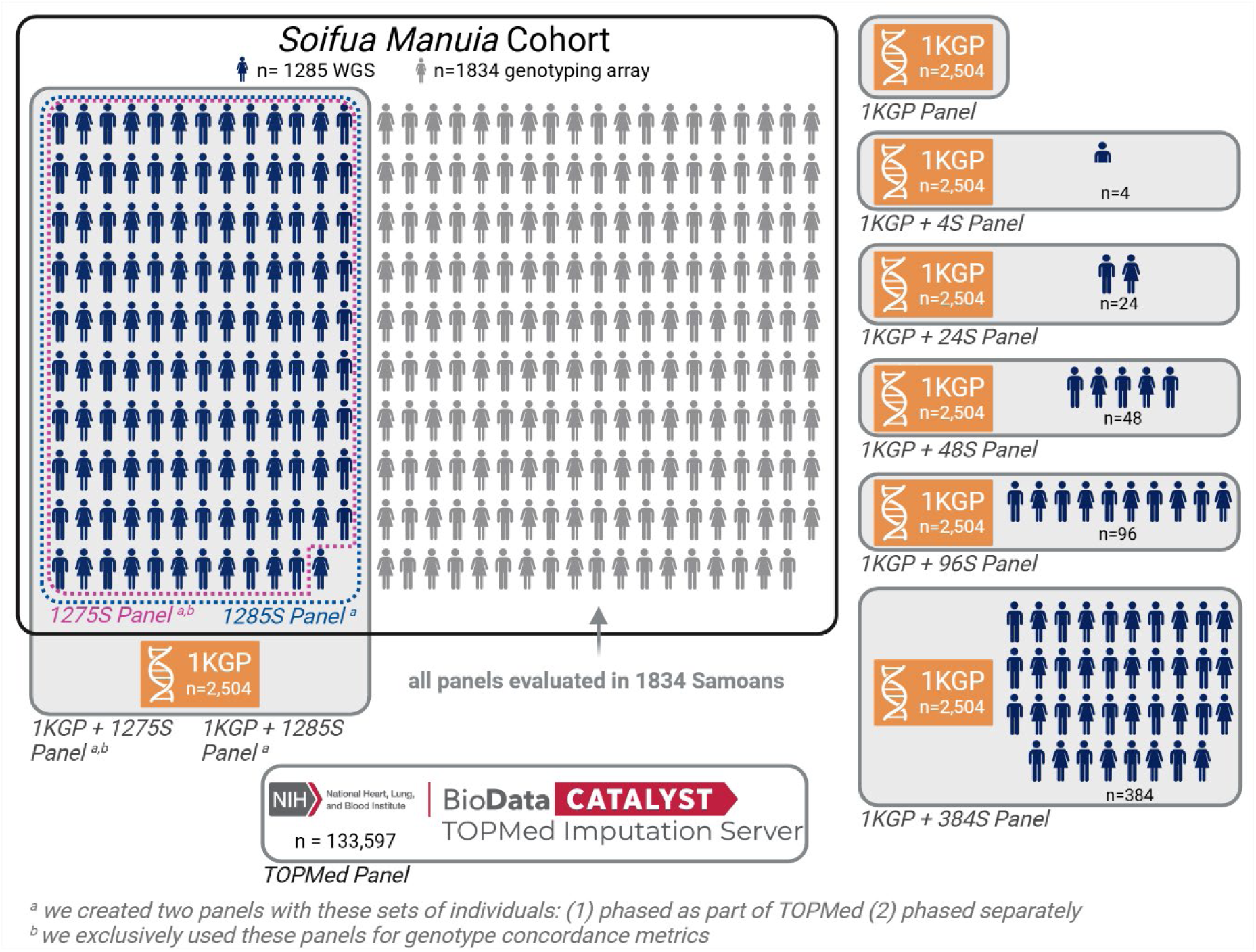
Graphical Depiction of the Reference Panels. The construction of the reference panels from Table 1 is represented below. Each person icon represents approximately 10 participants. Dark blue icons indicate participants with whole-genome sequencing, gray icons indicate those with genotyping array data only, and orange indicates the 1000 Genomes Project (1KGP) participants sequenced with TOPMed freeze 9. Panels indicated with an *a* were created in two ways: one from deriving the phased haplotypes from the jointly phased haplotypes of TOPMed freeze 9b and one from phasing the individuals in the panel separately. Panels indicated with a *b* were only used to calculate genotype concordance. Figure created with BioRender.com.

**Table 1.** Names and descriptions of tested imputation protocols. Protocols 1-4 were used to compare the newly constructed *Soifua Manuia* panel to TOPMed-R3, 1000 Genomes (1KGP) and to a Samoan only panel (1285S). The relative gain in imputation quality for additional Samoan sequences added to the panel was measured with protocols 5-9. Protocols 10-11 were used to directly measure genotype concordance with 10 hold out participants. Protocols 12-15 were compared with 11, 4, 10, and 2 to assess the impact of the phasing of the reference panel. Reference panel haplotypes were either extracted from TOPMed files (phased by TOPMed) or through phasing of the whole-genome sequences separately (phased in-house).

| Protocol | Name | Server | Individuals | Phased by TOPMed | Phased in-house |
| --- | --- | --- | --- | --- | --- |
| 1 | TOPMed-R3 | TOPMed Imputation Server | 133,597 TOPMed participants (no Samoans) | X |  |
| 2 | The *Soifua Manuia* panel (1KGP + 1285S) | In-house | 2,504 1KGP + 1285 Samoans | X |  |
| 3 | 1KGP | In-house | 2,504 1KGP (no Samoans) | X |  |
| 4 | 1285S | In-house | 1285 Samoans | X |  |
| 5 | 1KGP + 4S | In-house | 2,504 1KGP + 4 Samoans | X |  |
| 6 | 1KGP + 24S | In-house | 2,504 1KGP + 24 Samoans | X |  |
| 7 | 1KGP + 48S | In-house | 2,504 1KGP + 48 Samoans | X |  |
| 8 | 1KGP + 96S | In-house | 2,504 1KGP + 96 Samoans | X |  |
| 9 | 1KGP + 384S | In-house | 2,504 1KGP + 384 Samoans | X |  |
| 10 | 1KGP + 1275S | In-house | 2,504 1KGP + 1275 Samoans | X |  |
| 11 | 1275S | In-house | 1275 Samoans | X |  |
| 12 | 1275S in-house phasing | In-house | 1275 Samoans |  | X |
| 13 | 1285S in-house phasing | In-house | 1285 Samoans |  | X |
| 14 | 1KGP + 1275S in-house phasing | In-house | 2,504 1KGP + 1275 Samoans |  | X |
| 15 | 1KGP + 1285S in-house phasing | In-house | 2,504 1KGP + 1285 Samoans |  | X |

We created the *Soifua Manuia* reference panel by combining 2,504 1KGP individuals with all 1,285 Samoan participants (protocol 2). For comparison, we created panels with only 1KGP (protocol 3) and only Samoan participants (protocol 4).

To assess the relative gain from having additional population-specific participants in the reference panel, we created panels by combining 1KGP with varying numbers of Samoan participants: 4, 24, 48, 96, and 384 (representing fractions/multiples of a 96-well plate; protocols 5-9). These participants were the first 4, 24, 48, 96, or 384 individuals based on the INFOSTIP ranking so that the reference panels reflected the most Samoan genetic diversity possible at that sample size (Figure S2).

For calculating genotype concordance among the panels with the most Samoan representation, we constructed panels with 1275 Samoan participants (protocols 10 and 11, with and without 1KGP respectively), holding out 10 participants distributed uniformly across the INFOSTIP rankings of individuals (ranks 120, 240, 360, 480, 600, 720, 840, 960, 1080, and 1200, chosen for generalizability and to specifically compare the performance of the panels with <100 Samoans).

All the haplotypes from panels described above were phased in a population-unaware manner by the TOPMed Informatics Research Center^3^, along with all other individuals present in freeze 9b. For protocols 12-15, we phased the sequence data comprising the reference panels separately using Eagle v2.4.1^34^, such that protocol pairs 1 and 15, 4 and 13, 10 and 14, and 11 and 12 only differ from each other in the way the reference panel was phased.

### Imputation

We imputed genotypes using all references panels into Samoans with Affymetrix genotyping array genotypes as the target. We imputed genotypes on chromosomes 21 (for computationally efficiency) and 5 (to measure imputation in a Samoan-specific variant of interest—rs200884524, a stop-gain variant associated with lipid levels in a prior study^31^) for all Samoan participants in the 2010 cohort who were not part of any reference panel (*n* = 1,834). To ensure that conclusions drawn from these two chromosomes did not misrepresent genome-wide trends, we also imputed genotypes on all autosomes for 100 randomly selected participants.

Prior to imputation, Affymetrix genotyped variant coordinates were converted to the hg38 genome build using LiftOver (https://genome.ucsc.edu/cgi-bin/hgLiftOver). Then, data were aligned to the reference panel using Genotype Harmonizer^35^, using the mafAlign option to align to the minor allele when LD alignment failed and the minor allele frequency (MAF) was ≤ 30% in both the input and reference sets. The resulting variants were then phased with Eagle v2.4.1^34^ and genome-wide imputation was performed using Minimac4^36^. An example of the imputation code is available at https://github.com/jennaccarlson/imputation.

To benchmark the performance of the in-house panels, we imputed variants in the same sets of Samoans using the TOPMed-R3 reference panel via the TOPMed Imputation Server^3^ (protocol 1). We also performed meta imputation between the TOPMed-R3 and 1285S imputed variants (protocols 1 and 4) with MetaMinimac2^37^ (https://github.com/yukt/MetaMinimac2). Importantly, MetaMinimac2 requires that the genotyping array data be phased consistently prior to meta imputation. Thus, we pre-phased the genotypes of the *n*=1834 Samoan participants with the TOPMed Imputation Server prior to imputing with our in-house 1285S reference panel for the meta-imputation comparison only.

We extracted imputation metrics from the resulting info files and tabulated the number of well-imputed (r^2^ ≥ 0.80), non-monomorphic variants for each imputation. For each variant, we predicted functional consequence and impact using VEP,^38^ and calculated enrichment for each variant consequence and SnpEff impact group^39^ for variants that were uniquely imputed in a Samoan panel (r^2^ ≥ 0.80 in either 1285S or 1KGP+1285S and r^2^ < 0.80 or absent from both TOPMed-R3 and 1KGP) compared to shared variants (r^2^ ≥ 0.80 in either 1285S or 1KGP+1285S and either TOPMed-R3 or 1KGP). Consequence and impact groupings are listed in Table S3. Enrichment p-values were calculated with a two-sided z test for the proportion of variants with the given annotation within the MAF stratum.

Additionally, we merged imputed variants by position and alleles, accounting for possible reference and alternate allele flips, and calculated mean and median r^2^ for the intersection of variants across MAF bins. We examined empirical r^2^ for the genotyped variants. We also assessed the impact of the reference panel to impute rs200884524, a population-specific variant of interest mentioned above, by examining imputed MAF and r^2^.

To assess if the imputation quality gains from using a reference panel that includes Samoan haplotypes were due to allele frequency differences or haplotype size, we calculated fixation index (F_ST_) using the Weir and Cockerham formula and LD score. F_ST_ was calculated using the *Soifua Manuia* panel (protocol 1) with vcftools^40^; the population labels used were derived from the super populations listed in 1KGP (AFR, AMR, EAS, EUR, SAS)^1^, with the 1,285 Samoans listed as a separate sixth population. LD scores were calculated using the *Soifua Manuia* reference panel (protocol 1) in GTCA^41^ with the default block size (1 Mb with an overlap of 500 kb between blocks). For visualization, F_ST_ and LD scores were each stratified into low and high at the median value within a given MAF bin (Table S8).

Additionally, we compared genotype concordance for 10 hold-out participants with WGS from imputations using the following reference panels: TOPMed-R3, 1KGP, 1KGP + 4S, 1KGP + 24S, 1KGP + 48S, 1KGP + 96S, 1KGP + 1275S, 1KGP + 1275S in-house phasing, 1275S, and 1275S in-house phasing (protocols 2, 3, 5-11). We calculated non-reference discordance for each of these panels’ imputed genotypes for the 10 hold-out participants with WGS on chromosome 21 using bcftools^42^.

We also investigated if the phasing of the reference panel impacted the imputation of population-specific variation. We hypothesized that for Samoan-specific variation, the phasing of genotypes when done alongside super majority populations with very different allele frequencies might be biased toward the haplotypes not common in Samoans, thereby decreasing the Samoan-specific allele frequency. To investigate this, we compared imputation quality (r^2^), MAF, and non-reference discordance for two sets of imputed, Samoan-specific variants, defined as those in the 99^th^ percentile of F_ST_ and with MAF > 0.05.

## Results

We compared the average imputation quality (r^2^), genotype concordance, and the number of well-imputed variants (r^2^ ≥ 0.8) across 11 sets of imputed genotypes to assess the performance of the various imputation reference panels which included TOPMed-R3, 1KGP, 2 panels of 1285 Samoan participants, and 8 panels constructed by combining 1KGP with varying numbers of Samoans (Table 1).

### Panels with more than 24 Samoans yield more well-imputed variants than TOPMed

Panels with 1285 Samoan participants (1KGP + 1285S and 1285S) resulted in the largest number of well-imputed variants, yielding ∼300,000 more variants on chromosome 5 and ∼70,000 more on chromosome 21 than the TOPMed-R3 panel, with over 25,000 and ∼6,500 of these having a MAF of at least 0.05 on chromosomes 5 and 21, respectively (Figure 2; Table S1). The yield in well-imputed variants increased as the number of Samoans included in the reference panel increased across all MAF bins (up to a 55% increase in the total number of well-imputed variants compared to the TOPMed-R3 imputation). The largest gains were seen for variants with MAF < 0.05, with up to a 187% increase in well-imputed variants compared to the TOPMed-R3 imputation. Moreover, the panels with the most Samoans (1KGP + 1285S and 1285S panels) had over 200,000 and 60,000 well-imputed unique variants (not seen in TOPMed-R3 or 1KGP imputations) on chromosomes 5 and 21, respectively, in addition to the variants shared across panels (Figure 3). We annotated these shared and unique variants for consequence and impact and observed enrichment in several categories among our uniquely imputed variants, including significantly more modifier, moderate, and high impact variants (Figure 4, Table S4-S5).

**Figure 2.**
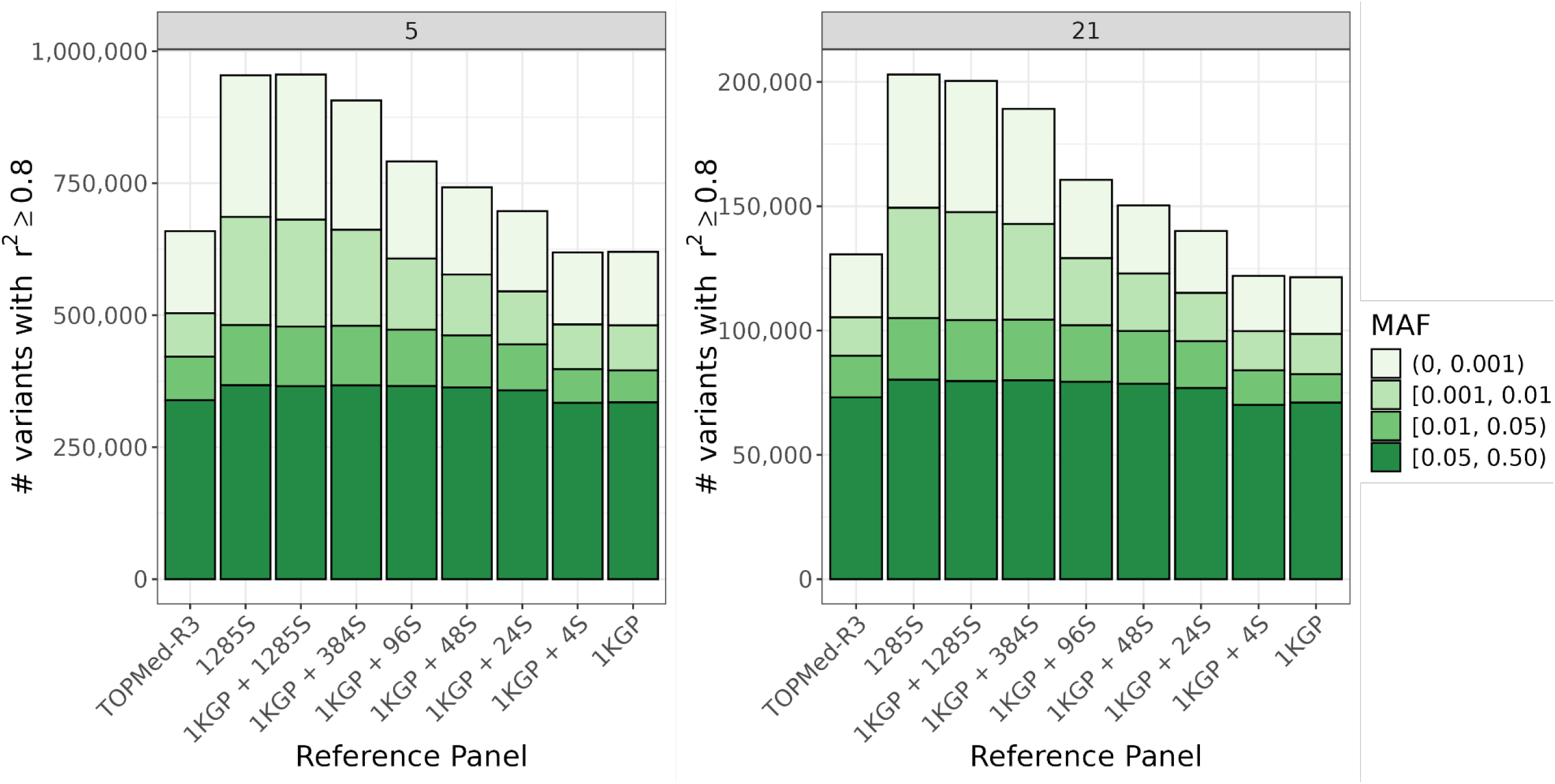
Count of well-imputed variants by reference panel. The number of variants with imputation quality r^2^ ≥ 0.80 by reference panel is plotted across minor allele frequency (MAF) bins for chromosomes 5 and 21. (These counts are printed in Table S1.)

**Figure 3.**
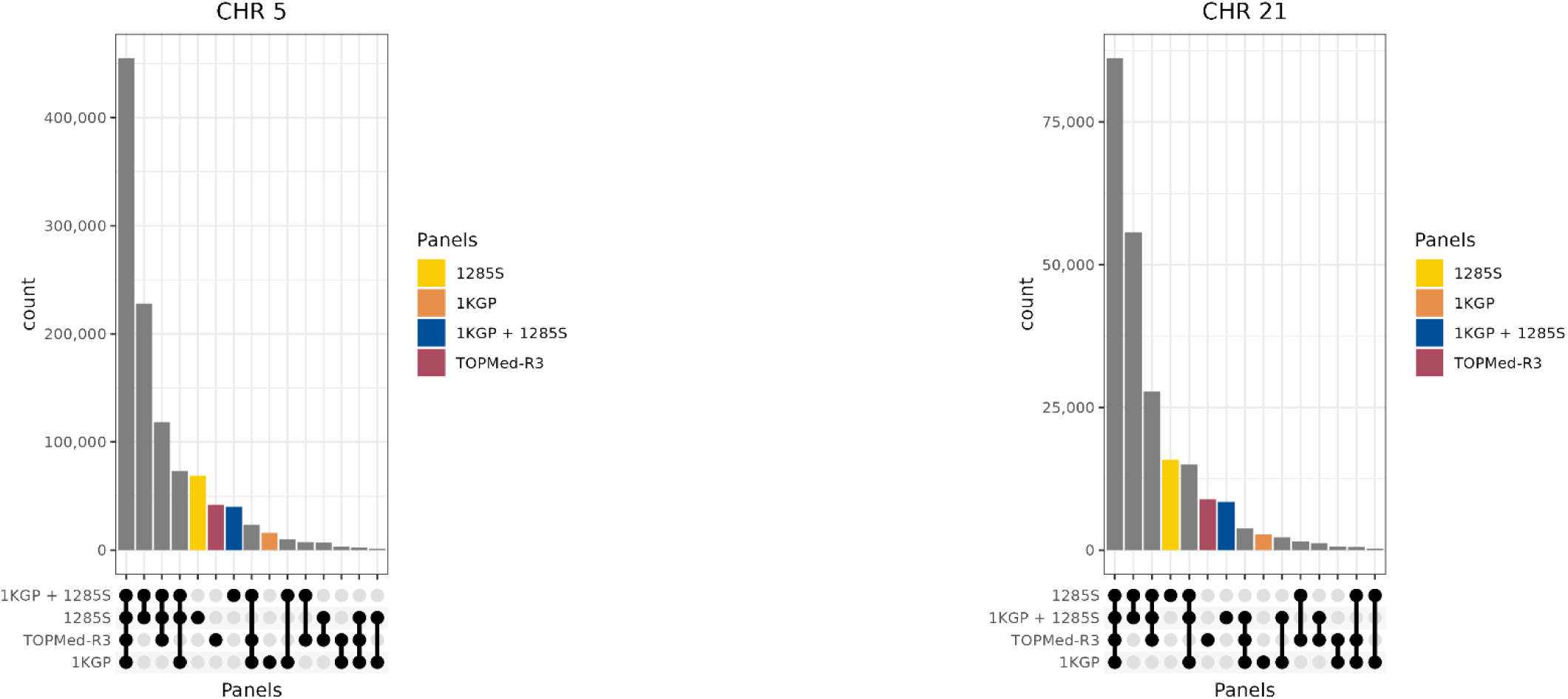
Count of uniquely imputed variants by combinations of reference panels. The number of well imputed variants (r^2^ ≥ 0.80) is plotted for chromosomes 5 and 21 across combinations of four reference panels (1285S, 1KGP + 1285S, TOPMed-R3, and 1KGP) to demonstrate how many variants are imputed similarly or uniquely across the panels. Black dots in the bottom panels indicate which reference panels contained the variants. For example, the first bar represents variants with r^2^ ≥ 0.80 across all four panels, while the second bar represents those with r^2^ ≥ 0.80 only in 1KGP + 1285S and 1285S.

**Figure 4.**
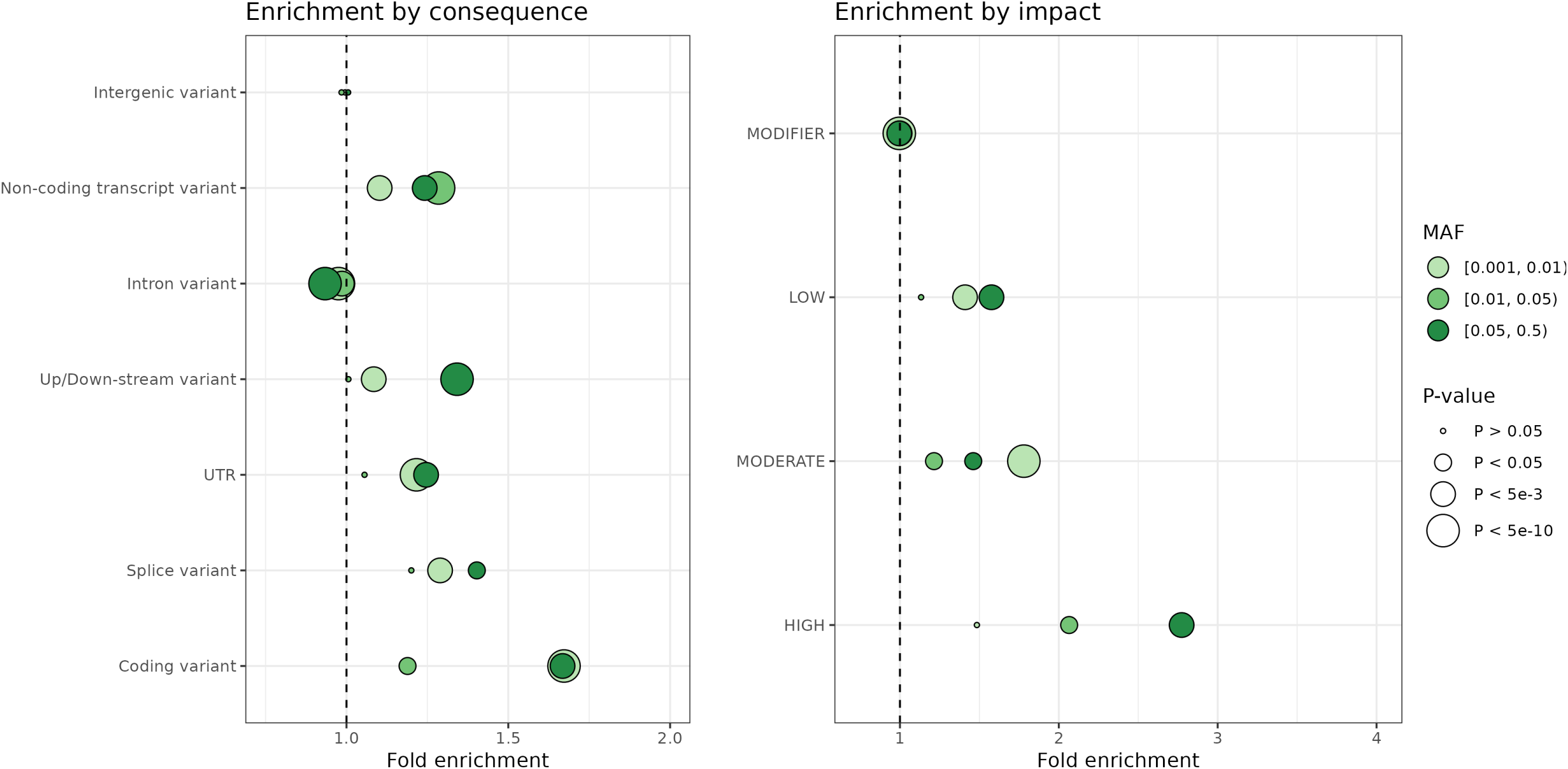
Enrichment of variants from Samoan imputation. For well-imputed (r^2^ ≥ 0.80) variants on chromosomes 5 and 21, variants that were unique (present in 1285S and/or 1KGP + 1285S and not in TOPMed-R3 nor 1KGP) and shared (present in 1285S or 1KGP + 1285S and TOPMed-R3 or 1KGP) were annotated by consequence and impact with VEP. Enrichment was calculated by comparing the proportion of variants in that grouping and MAF bin between unique and shared variants so that a fold enrichment > 1 indicates a variant type more common in the uniquely imputed variants. The color of points indicates MAF bin, and the size of points scales with p-value.

We also examined “lost” variants, those with imputation quality r^2^ < 0.80 in the 1KGP imputation, to see how many could be “rescued” (had imputation quality r^2^ ≥ 0.80) across other panels (Figure 5; Table S2). The proportion of lost variants rescued was highest for variants with greater MAF across all panels, with nearly 100% of lost variants recovered in some cases (MAF ≥ 0.05 for the 1KGP + 1285S panel). The panels with at least 48 Samoans included rescued more variants overall than TOPMed-R3 (15.0% to 35.5% for Samoan panels compared to 12.2% to 13.2% for TOPMed). Over 95% of lost variants with MAF ≥ 0.05 were rescued for the reference panels with 384 or more Samoans. Even for the 1KGP + 4S panel, 29.4% to 37.1% of variants with MAF ≥ 0.05 were rescued.

**Figure 5.**
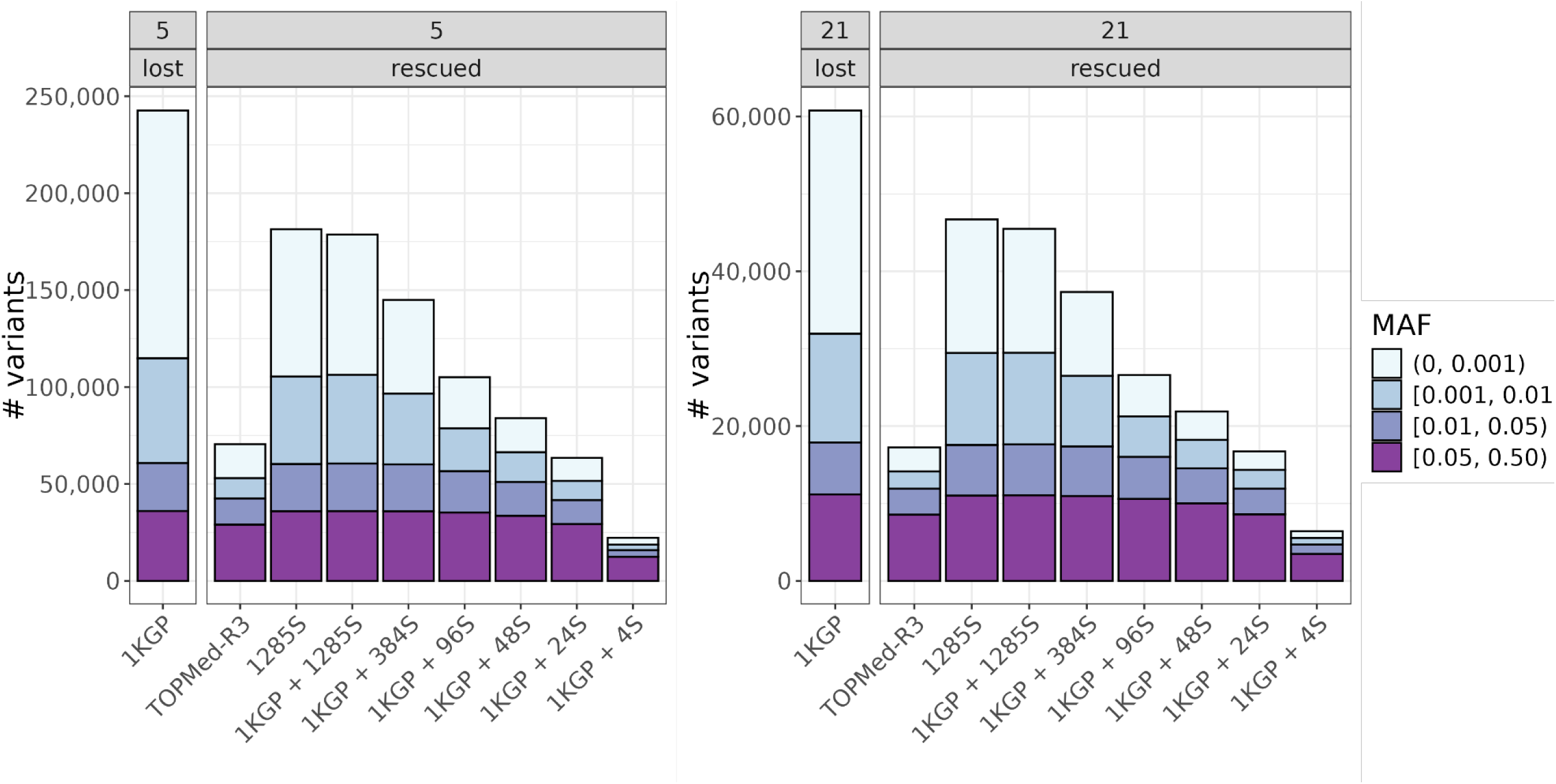
Count of rescued variants by reference panel. The number of imputed variants that were lost (r^2^ < 0.80) in the 1KGP imputation but were rescued (r^2^ ≥ 0.80) in other panels by minor allele frequency (MAF) and reference panels across chromosomes.

### Similar imputation quality to TOPMed is achieved with as few as 24 Samoans

Imputation quality increased as the number of Samoans represented in the panel increased. The panels with 48 or more Samoans included outperformed the TOPMed-R3 panel for all variants with imputed MAF ≥ 0.001 (based on the 1KGP + 1285S imputation), while the 1KGP+24S performed similarly to TOPMed-R3 (Figure 6; Tables S6-S7). When looking at empirical r^2^ for genotyped variants only, which is a more robust metric for evaluating imputation for lower frequency variants, the panels including at least 24 Samoans outperform both the TOPMed-R3 and 1KGP panels (Figure 7). This trend was consistent across all autosomes (Figure S9). Average imputation quality tended to be lower when examining variants based on TOPMed-imputed MAF instead of 1KGP + 1285S-imputed MAF (Figure 6), consistent with expectations that Samoan allele frequencies would differ from those in TOPMed-R3 due to lower-quality matches of local haplotypes in the TOPMed-R3 reference panel. However, even when using TOPMed-imputed MAF, the panels containing 24 or more Samoans yielded the same or higher quality compared to TOPMed-R3 panel across the allele frequency spectrum. Consistent with this, the non-reference discordance for TOPMed-R3 (4.44) was higher than that for 1KGP + 24S (4.19), 1KGP + 48S (3.14), 1KGP + 96 (2.43), 1KGP + 1275S (0.81), and 1275S (0.84) panels (Table 2). Meta-imputation between the 1275S and TOPMed-R3 results yielded higher discordance (2.22) than the Samoan only panel alone, but lower discordance than the TOPMed-R3 panel alone (Table 2).

**Figure 6.**
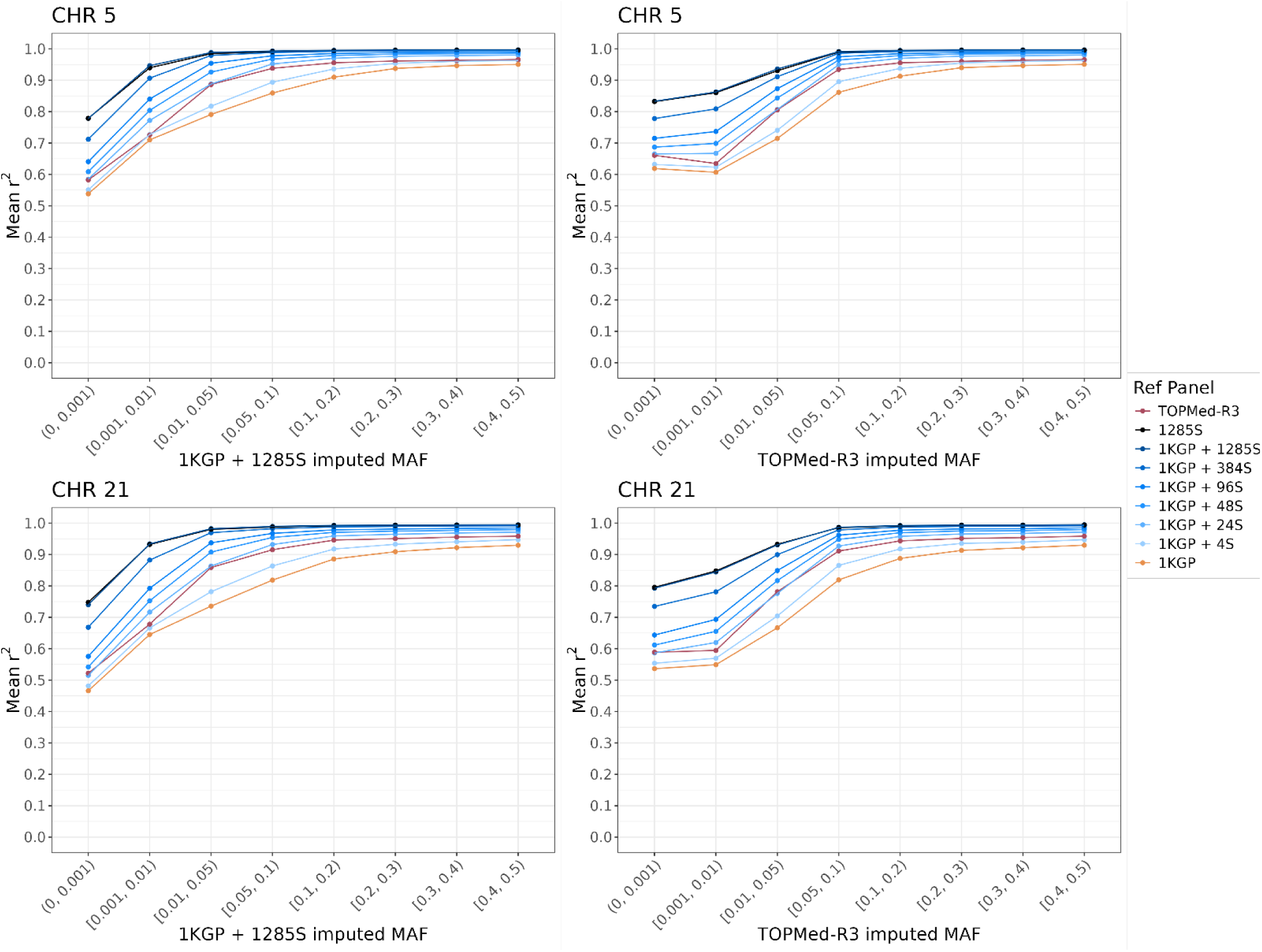
Imputation quality versus minor allele frequency by reference panel. Mean imputation quality (r^2^) by minor allele frequency (MAF) bin (left column: MAF based on 1KGP + 1285S imputation; right column: MAF based on the TOPMed-R3 imputation) is plotted by reference panel on chromosomes 5 and 21. (Plots with median r^2^ are given in Figure S3. Tables with mean and median r^2^ across reference panel are in Table S6-S7.)

**Figure 7.**
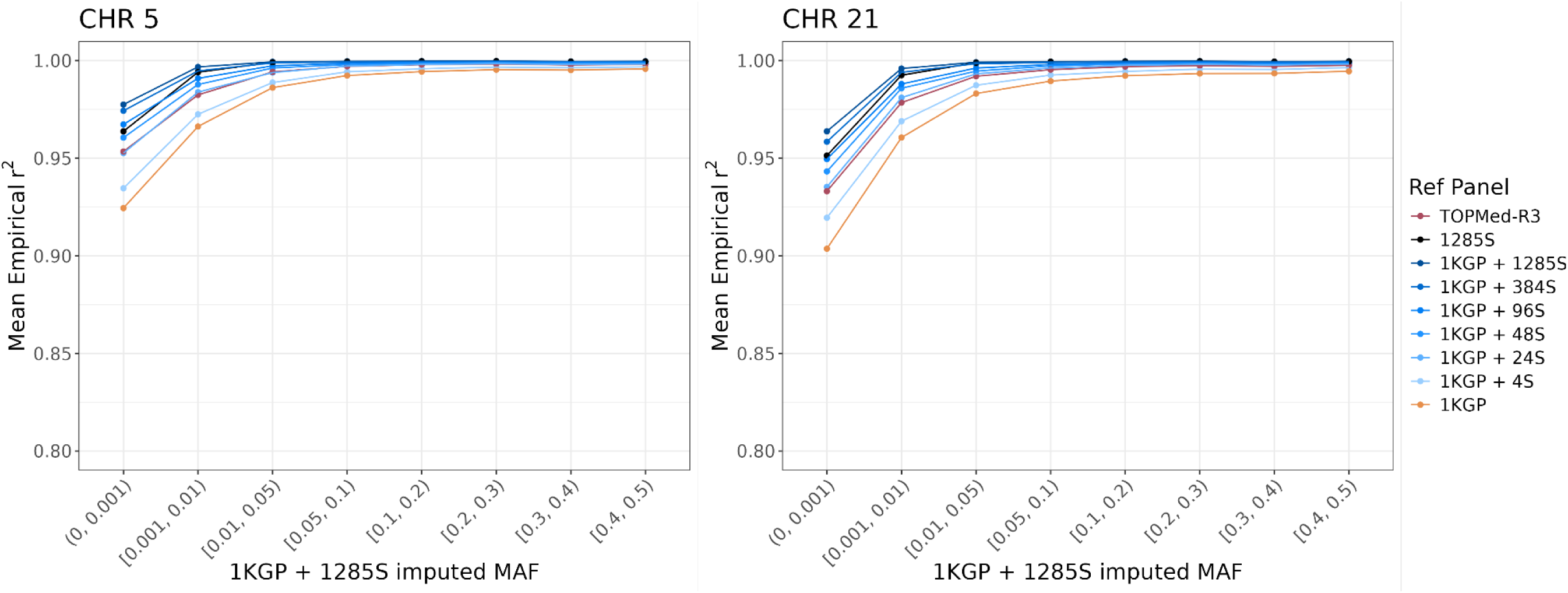
Empirical r^2^ versus minor allele frequency by reference panel for genotyped variants. Mean empirical r^2^ by minor allele frequency (MAF) bin (based on 1KGP + 1285S imputation) is plotted by reference panel on chromosomes 5 and 21. Y-axis limited to 0.8-1.0 to show differences across panels. (Plots with median empirical r^2^ are given in Figure S4.)

**Table 2.** Non-reference discordance on chromosome 21 calculated from 10 hold-out participants with WGS. Discordance was calculated for two sets of variants: all single nucleotide variants on the chromosome (SNVs) and 598 Samoan-specific SNVs (defined as being in the 99^th^ percentile of F_ST_ and having MAF ≥ 0.05 in Samoans). The 10 hold-out participants were chosen based on INFOSTIP ranking (ranks 120, 240, 360, 480, 600, 720, 840, 960, 1080, and 1200) to capture a range of haplotypes present in sample and facilitate the evaluation of the panels with <100 Samoans.

| Protocol | Reference Panel | Non-Reference Discordance |  |
| --- | --- | --- | --- |
|  |  | All SNVs | 598 Samoan-specific SNVs |
| 1 | TOPMed-R3 | 4.44 | 3.17 |
| 3 | 1KGP | 10.21 | 4.34 |
| 5 | 1KGP + 4S | 7.66 | 3.18 |
| 6 | 1KGP + 24S | 4.19 | 1.87 |
| 7 | 1KGP + 48S | 3.14 | 1.51 |
| 8 | 1KGP + 96S | 2.43 | 1.40 |
| 10 | 1KGP + 1275S | 0.81 | 0.61 |
| 14 | 1KGP + 1275S in-house phasing | 0.99 | 0.59 |
| 11 | 1275S | 0.84 | 0.56 |
| 12 | 1275S in-house phasing | 0.99 | 0.62 |

### Gains in imputation quality from Samoan representation in the reference panel are greatest for high F_ST_ variants and low LD score variants

Imputation quality generally increased for all panels as F_ST_ increased, as expected due to higher confidence in matching haplotypes with population-specific variation (Figure 8). However, for the highest three F_ST_ deciles, the mean r^2^ for TOPMed-R3 panel dipped slightly. We also stratified F_ST_ into high and low strata about the median value within each MAF bin to examine the relative difference in imputation quality by F_ST_ (Table S8). For variants with MAF < 0.05, those with high F_ST_ (i.e., F_ST_ greater than the median F_ST_ of all variants within the same MAF bin) tended to be imputed better across all reference panels except the TOPMed-R3 panel, which had similar or slightly lower r^2^ for high F_ST_, low-frequency variants. For variants with MAF ≥ 0.05, there did not appear to be a substantial difference in imputation quality by F_ST_ except for the 1KGP panel (Figure 9). A similar trend was observed for LD scores, with imputation quality increasing as LD score increased (Figure 10) and high LD score variants (i.e., LD score greater than the median LD score of all variants within the same MAF bin (Table S8)) being imputed better on average than low LD score ones across the MAF spectrum, consistent with expectations (Figure 11).

**Figure 8.**
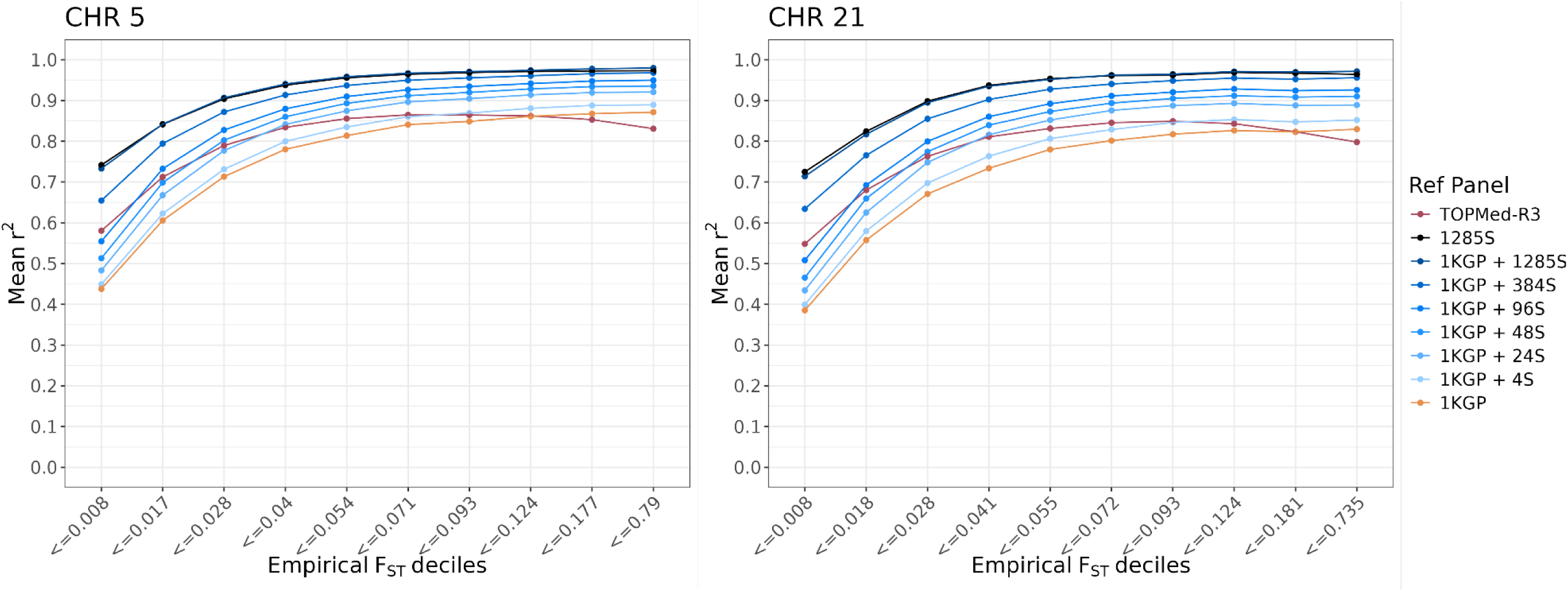
Imputation quality versus fixation index by reference panel. Mean imputation quality (r^2^) by empirical Fixation Index (F_ST_) deciles is plotted by reference panel on chromosomes 5 and 21. (Plots with median r^2^ are given in Figure S5.)

**Figure 9.**
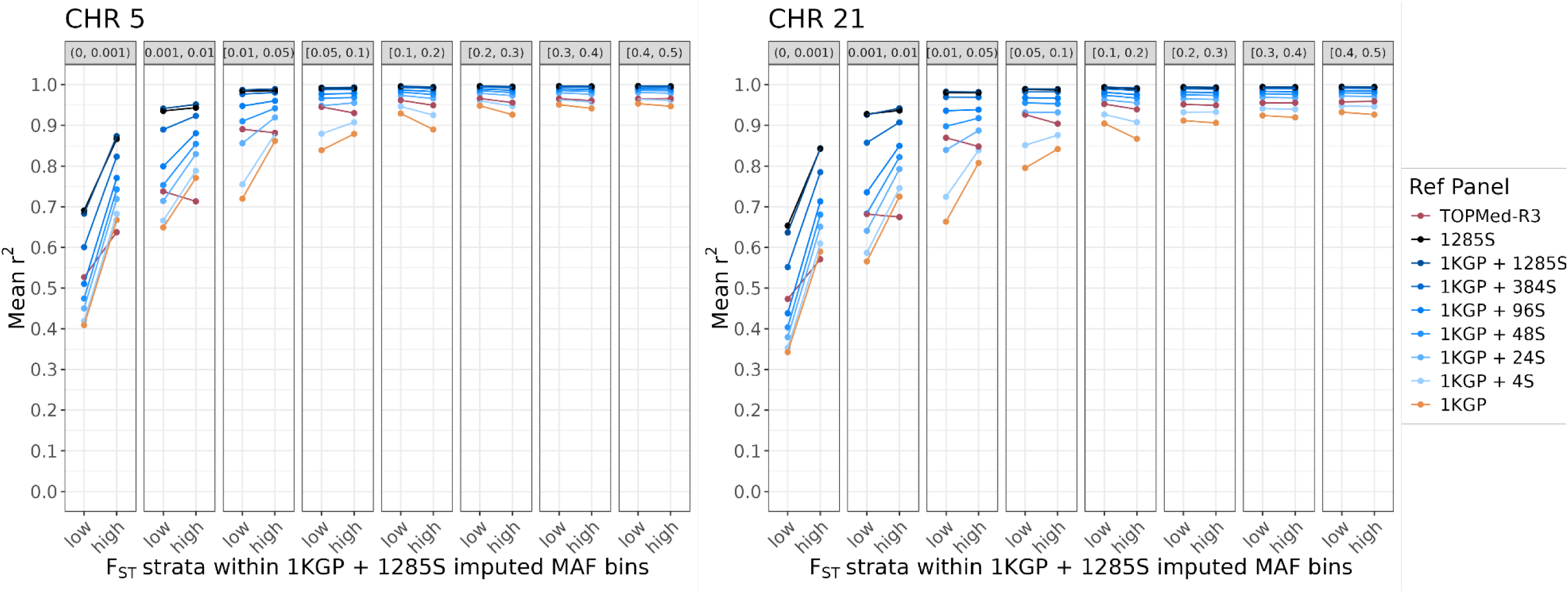
Imputation quality versus fixation index by reference panel and MAF bin. Mean imputation quality (r^2^) by median-stratified Fixation Index (F_ST_) across MAF (based on 1KGP + 1285S imputation) is plotted by reference panel on chromosomes 5 and 21. F_ST_ was stratified into ‘low’ and ‘high’ about the median F_ST_ within each MAF bin (values given in Table S8). (Plots with median r^2^ are given in Figure S6.)

**Figure 10.**
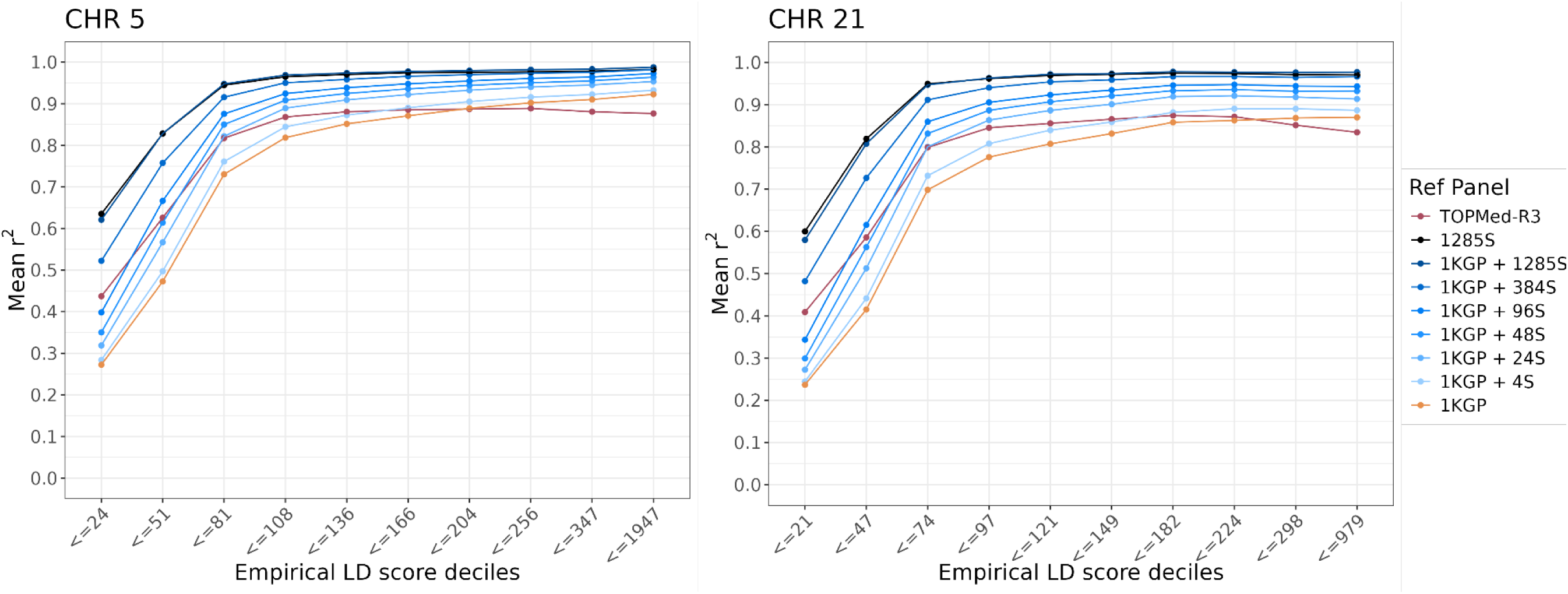
Imputation quality versus LD score by reference panel. Mean imputation quality (r^2^) by empirical LD score deciles is plotted by reference panel on chromosomes 5 and 21. (Plots with median r^2^ are given in Figure S7.)

**Figure 11.**
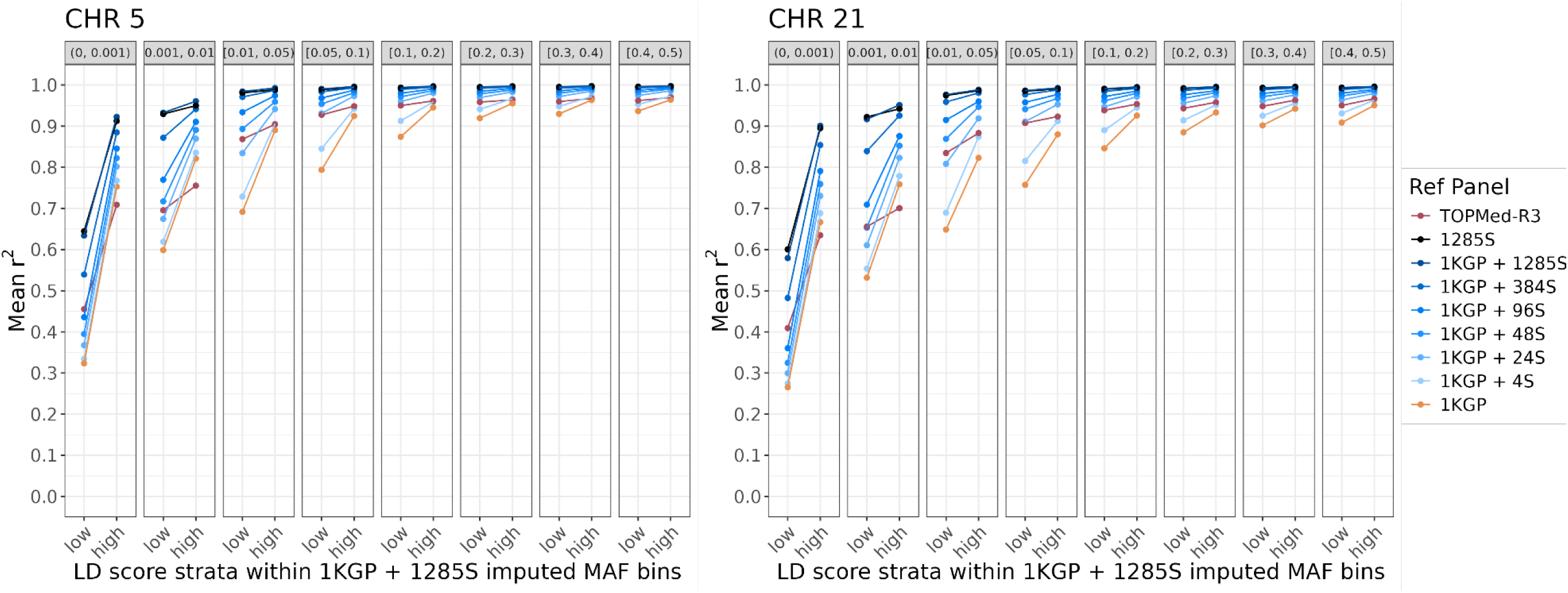
Imputation quality versus LD score by reference panel and MAF bin. Mean imputation quality (r^2^) by median-stratified LD score across MAF (based on 1KGP + 1285S imputation) is plotted by reference panel on chromosomes 5 and 21. LD score was stratified into ‘low’ and ‘high’ about the median LD score within each MAF bin (values given in Table S8). (Plots with median r^2^ are given in Figure S8.)

To identify which variants had higher imputation quality with the 1KGP + 1285S panel than with the TOPMed-R3 panel, we compared the difference in mean r^2^ across the two sets of imputed variants. The gains in average r^2^ for the 1KGP + 1285S panel compared to the TOPMed-R3 panel were more pronounced for lower frequency variants (0.001 ≤ MAF < 0.05) with low F_ST_ and low LD scores (Figure 12). However, for variants with MAF ≥ 0.05, there was no discernible difference in average imputation quality across low and high F_ST_ variants but a small difference comparing low and high LD score variants. Low LD score variants had a larger gain in imputation quality than low F_ST_ variants for the 1KGP + 1285S panel over the TOPMed-R3 panel. This suggests that the gains in imputation from the 1KGP + 1285S panel over TOPMed-R3 were not only from allele frequency differences but also from haplotype structure and that the imputation was the most improved for lower-frequency variants on smaller haplotypes.

**Figure 12.**
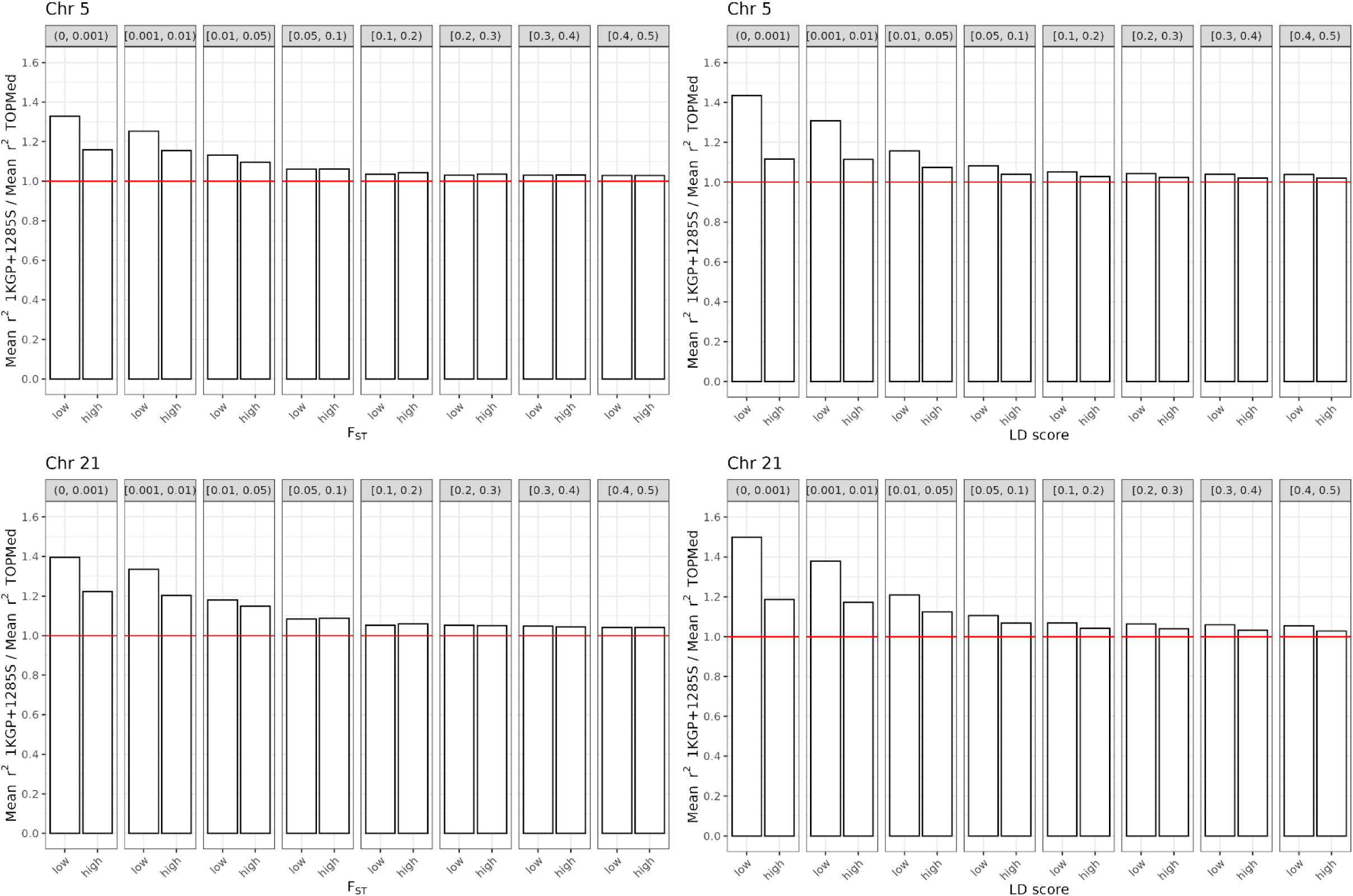
Gains in imputation quality from TOPMed-R3 panel to 1KGP + 1285S panel by MAF for F_ST_ and LD score. Fold change in mean imputation quality (r^2^ 1KGP+1285S / r^2^ TOPMed-R3) by median-stratified F_ST_ (left) and LD scores (right) across MAF (based on 1KGP + 1285S imputation) is plotted for chromosomes 5 and 21. Both F_ST_ and LD score were stratified into ‘low’ and ‘high’ about the median F_ST_/LD score within each MAF bin (values given in Table S5). Reference line at *y* = 1 (red) is given to indicate no difference between TOPMed-R3 and 1KGP + 1285S panels.

### Improvement in the imputation of population-specific variation

For rs200884524 the imputation quality is highest (r^2^ = 0.89-0.95) for the reference panels that include Samoan haplotypes (Table 3). Additionally, the MAF among the imputed genotypes in the 1,834 non-reference-panel Samoans from protocols 3-11 (0.204-0.222) is much closer to what is expected – targeted genotyping in separate cohorts of similar ancestry yielded a MAF of 0.202-0.233^31^. The TOPMed-R3 imputation yielded a much more accurate imputed MAF than 1KGP, which suggests the presence of some Pacific haplotypes at this locus in the TOPMed-R3 reference panel even though the T (minor) allele count of this variant is still low in TOPMed (55 alleles present out of 301,798 total alleles in TOPMed freeze 10 according to BRAVO^43^; https://bravo.sph.umich.edu/). Examining the genotype concordance of Samoan-specific variation (F_ST_ above the 99^th^ percentile and having MAF ≥0.05 in Samoans) yielded similar conclusions; panels with Samoans included had the lowest non-reference discordance for these variants and TOPMed-R3, while worse than Samoan panels with at least 24 Samoans, still outperformed 1KGP (Table 2).

**Table 3.** Imputation quality and minor allele frequency (MAF) for rs200884524 (chr5:181050285 C/T [hg38]) in *BTNL9* across reference panels. Targeted genotyping in two separate cohorts of 1,666 Samoan and American Samoan adults yielded a MAF of 0.202-0.233^31^.

| Protocol | Reference panel | $r^2$ | MAF |
| --- | --- | --- | --- |
| 1 | TOPMed-R3 | 0.84740 | 0.17385 |
| 3 | 1KGP | 0.45654 | 0.01361 |
| 5 | 1KGP + 4S | 0.90687 | 0.20424 |
| 6 | 1KGP + 24S | 0.91276 | 0.22198 |
| 7 | 1KGP + 48S | 0.86967 | 0.20785 |
| 8 | 1KGP + 96S | 0.91062 | 0.20710 |
| 9 | 1KGP + 384S | 0.94567 | 0.22130 |
| 2 | 1KGP + 1285S | 0.94360 | 0.21852 |
| 15 | 1KGP + 1285S in-house phasing | 0.94360 | 0.21852 |
| 4 | 1285S | 0.93441 | 0.21926 |
| 13 | 1285S in-house phasing | 0.93036 | 0.22106 |

### Phasing of the reference panel can affect imputation of population-specific variation

We compared r^2^ between variants imputed with reference panels that only differed in how they were phased (protocols 1 & 15, 4 & 13) for Samoan-specific variants (i.e., those in the 99^th^ percentile of F_ST_ that had MAF ≥ 0.05 in Samoans). For the panels comprised of 1KGP + 1285S there were no systematic differences in imputation quality by phasing of the reference panel.

The correlation between imputed r^2^ from the 1KGP +1285S panel and that from the 1KGP + 1285S in-house phasing panel for the 3,410 Samoan-specific variants was ∼0.999 (Figure 13, bottom row). However, for the panels comprising only the 1285 Samoans (without 1KGP), imputed r^2^ was lower for the panel that was phased in-house (Figure 12, top row), contrary to our hypothesis. The correlation between imputed r^2^ from the 1285S panel and that from the 1285S in-house phasing panel was lower than for the 1KGP + 1285S panels (r= ∼0.95). When examining all variants aggregated (not examining Samoan-specific variation), the panels that only differed in the way they were phased performed very similarly for imputation r^2^ (Tables S1, S2, S6, S7). Through examining genotype concordance, we observed that the panels that were phased in-house (with or without 1KGP) had slightly higher non-reference discordance for all variants (Table 2), suggesting that which additional participants are included in the phasing of the reference panel can impact imputation quality.

**Figure 13.**
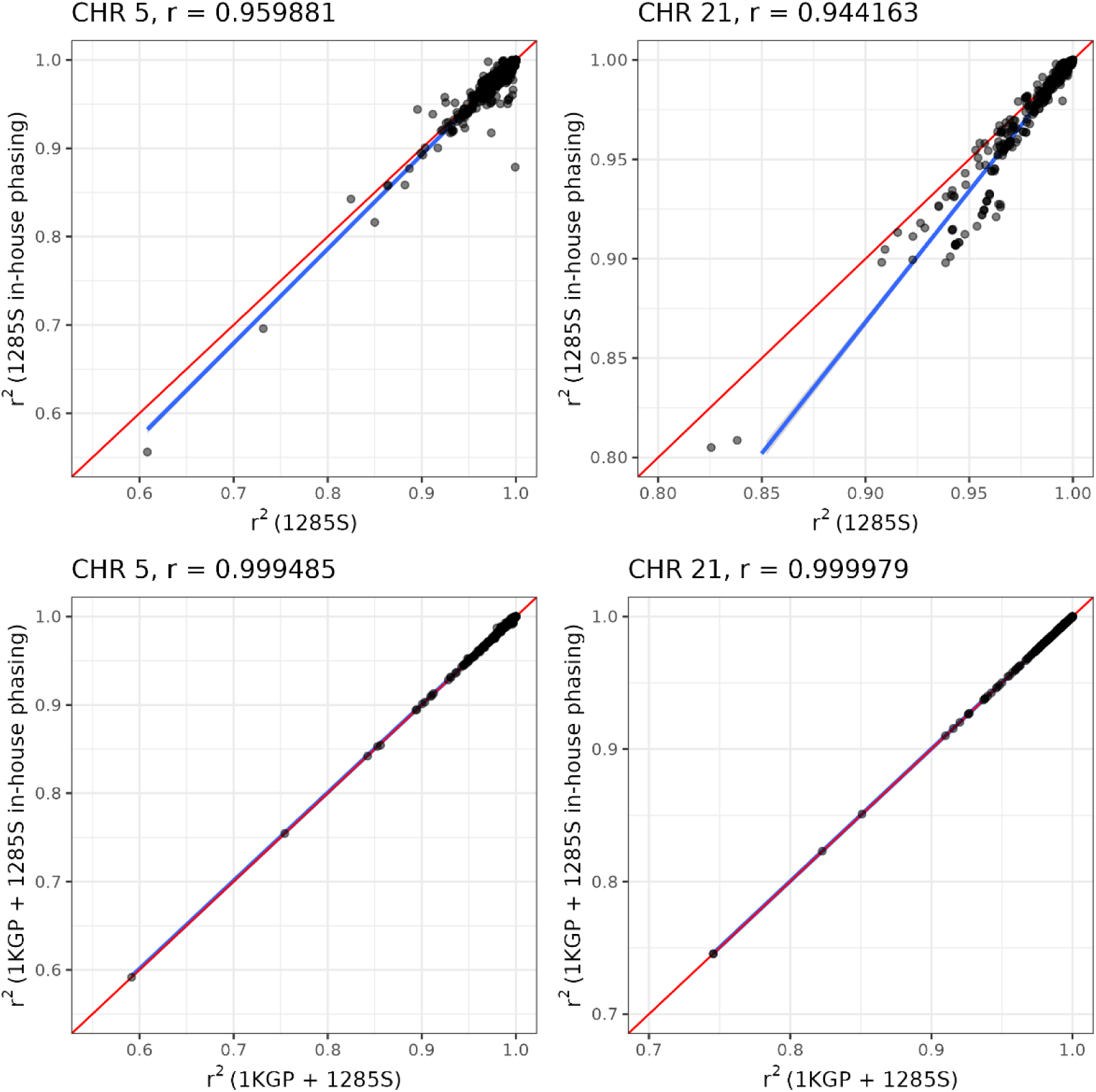
Comparison of imputation quality by phasing of the reference panel. Imputation quality (r^2^) for variants in the 99^th^ percentile of F_ST_ that were common in Samoans (MAF ≥ 0.05) across chromosome 5 and 21. Top row: the 1285S panel (X-axis; protocol 10) versus the 1285S in-house phasing panel (Y-axis; protocol 11). Bottom row: the 1KGP + 1285S panel (X-axis; protocol 8) versus the 1KGP + 1285S in-house phasing panel (Y-axis; protocol 9). Red line indicates Y=X line. Blue line is fitted linear regression line based on the observed r^2^ values. Correlation (r) between the imputed r^2^ from the two panels is given above each plot.

### Meta imputation has worse performance than the population-specific panel alone

We performed meta imputation of the 1285S panel with the TOPMed-R3. Prior to meta imputation, we phased the genotyping data of the *n*=1834 target participants with the TOPMed Imputation Server, which through its standard QC process to align the target genotypes to the TOPMed reference panel, resulted in 3,503 fewer variants in the target genotypes (2,840 and 663 on chromosomes 5 and 21, respectively).

The average imputation r^2^ for meta imputation fell in between that for the 1285S and TOPMed-R3 panels across the MAF spectrum (Figures S10, S12).

The meta imputation yielded the highest number of well-imputed variants (r^2^ ≥ 0.80) compared to the 1285S and TOPMed-R3 panels (Figure S11, Table S9), but fewer than the *Soifua Manuia* panel (Table S1). Notably, there was a difference in the number of well-imputed variants with the 1285S panel when using pre-phased target genotypes from the TOPMed Imputation Server (861,094 and 179,604 variants with r^2^ ≥ 0.8 on chromosomes 5 and 21, respectively) compared to those from the 1285S panel when phasing the target genotypes in-house (954,408 and 202,920 variants with r^2^ ≥ 0.8 on chromosomes 5 and 21, respectively).

## Discussion

The *Soifua Manuia* reference panel consisting of 1KGP + 1285 Samoans outperformed all other panels tested, especially for low-frequency variants. There were substantial gains in imputation quality (r^2^) and in the number of well-imputed variants across the allele frequency spectrum, with more pronounced gains for low-frequency variants. Compared to the commonly recommended TOPMed-R3 panel, the number of well-imputed variants available across chromosomes and MAF ranges increased by 8% to 182% with the 1KGP + 1285S panel and average imputation quality increased by 3% to 42%. When examining meta imputation between a population-specific panel (1285S) and the TOPMed-R3 panel, we observed lower imputation quality on average with meta imputation compared to the population specific panel, contrary to prior work demonstrating the superiority of meta imputation to single reference panel imputation^37,44^. This demonstrates that while TOPMed-R3 panel has improved imputation quality for many diverse populations, there are still populations that are not well represented by this or other publicly available panels.

Additionally, there was an enrichment in high impact, coding variants among those that were uniquely well-imputed by a Samoan panel (1285S or 1KGP + 1285S) compared to TOPMed-R3 and 1KGP. This has important implications for genetic association testing and fine-mapping of GWAS loci as measuring association with these variants that likely impact gene function would elucidate the putative causal gene, as we have previously demonstrated with the discovery of the stop-gain variant rs200884524^31^. As a proof of concept, we explicitly examined this variant and demonstrated that the imputation quality and minor allele frequency changed across the tested reference panels, with the most accurate imputation occurring with the *Soifua Manuia* panel. This has implications for post-imputation association testing for similar population-specific variants as they may be poorly imputed or imputed with less non-reference genotypes (i.e., with lower MAF) by panels not including population-specific haplotypes.

In looking at variants with low imputation quality in the 1KGP imputation, a large proportion of them were “rescued”, i.e., had high imputation quality, in panels containing 48 or more Samoans. These panels rescued more variants, especially common variants, than the TOPMed-R3 panel did. Across the variant frequency spectrum, with only 24 Samoans, we achieved the same imputation performance as TOPMed-R3 and had greater performance with all panels with 48 or more Samoans. Importantly, this sample size is not necessarily prescriptive for future studies planning to perform WGS to create their own population-specific as these 48 individuals were not randomly selected—they were chosen to maximize the genetic variation present in the original set of ∼3,000 participants, which were all of Samoan ethnicity (defined by participant self-reporting four Samoan grandparents) and therefore comprised a quite homogeneous population with little to no admixture^8^. These findings may reflect the fact that in this cohort, the 48 individuals yielded over 70% of the information content from INFOSTIP (Figure S2). In other populations, the number of individuals needed to see improved imputation will depend on the genetic variation of the target population. For populations with larger genetic heterogeneity, including those with lots of admixture or different haplotype sizes, more individuals will likely be needed to achieve improved imputation accuracy over extant panels.

The trends in imputation quality by F_ST_ and LD score suggest that these improvements in r^2^ for the *Soifua Manuia* panel are driven by differences in both the LD patterns and haplotypes of Samoans and allele frequency differences across populations. While imputation quality was highest for high LD score variants, the gains in imputation quality for the *Soifua Manuia* panel over the TOPMed-R3 panel were the largest for low LD score variants across the MAF spectrum. This suggests that the gains in imputation quality for the *Soifua Manuia* panel over TOPMed-R3 is driven by population-specific variants in small LD blocks that would not be predicted well by other reference panels.

We hypothesized that the phasing of reference panel haplotypes would affect imputation quality, specifically, that phasing taking place in large multiethnic consortia might bias the phasing of the reference panel haplotypes more common in majority-represented populations. In this study we observed an opposite effect to what we hypothesized—the imputation quality was slightly lower for population specific variation within a population-specific panel (1285S) phased separately. We did not observe this trend with the cosmopolitan panel (1KGP + 1285S). We also observed differences in genotype concordance by phasing of the reference panel – with those having been phased separately (1285S in-house phasing and 1KGP + 1285S in-house phasing) having slightly higher discordance than those phased with other studies as part of a consortium (1285S and 1KGP + 1285S). Based on our findings in this study, it does not appear that separate phasing yields better imputation.

Taken together, this work highlights the utility of augmenting cosmopolitan extant panels with population-specific sequences, resulting in more well-imputed variants for association testing, higher imputation quality overall, and better imputation of population-specific variation. In this work, these gains were the greatest for low-frequency population-specific variants in smaller LD blocks. These results suggest that adding even a small number of population-specific haplotypes can substantially increase the number of well-imputed variants available for analysis compared to gold-standard extant panels, which may allow for better inclusion of even more diverse populations in genetic association studies. This improvement in the representativeness of genetic studies, by extension, will lay the foundation for more equitable access to personalized medicine innovation in this and other Pacific populations.

## Data Availability

Upon publication, the Minimac files for the Soifua Manuia reference panel will be made available through dbGaP (accession number: phs000972.v5.p1). The discovery cohort data used for this study are available through dbGaP (accession numbers: phs000914.v1.p1 and phs000972.v5.p1). A template for the imputation code is available at https://github.com/jennaccarlson/imputation; additional code for creating the imputation reference panels and imputation quality analysis will be made available upon request.

## Acknowledgements

We would like to thank the Samoan participants of the study, local village authorities, and the many Samoan and other field workers over the years. We acknowledge the Samoan Ministry of Health, the Samoa Bureau of Statistics, and the Samoan Ministry of Women, Community and Social Development for their support of this research. We give particular thanks to two research assistants, Melania Selu and Vaimoana Lupematasila, who contributed to the 2010 recruitment and continue to assist us in our work in Samoa.

Molecular data for the Trans-Omics in Precision Medicine (TOPMed) program was supported by the National Heart, Lung, and Blood Institute (NHLBI). Genome Sequencing for NHLBI TOPMed: Samoan (phs000972.v5.p1) was performed at New York Genome Center Genomics (HHSN268201500016C) and Northwest Genomics Center (HHSN268201100037C). Core support including centralized genomic read mapping and genotype calling, along with variant quality metrics and filtering were provided by the TOPMed Informatics Research Center (3R01HL117626-02S1; contract HHSN268201800002I). Core support including phenotype harmonization, data management, sample-identity QC, and general program coordination were provided by the TOPMed Data Coordinating Center (R01HL120393; U01HL120393; contract HHSN268201800001I). We gratefully acknowledge the studies and participants who provided biological samples and data for TOPMed.

This work was funded by the National Institute of Health grants R01HL093093 (STM), R01HL133040 (RLM), R01AG009375 (STM), R01HL052611 (MI Kamboh), R01DK059642 (STM), and R01DK055406 (RD). Genotyping was performed in the Core Genotyping Laboratory at the University of Cincinnati, funded by National Institutes of Health grant P30ES006096 (S Pinney).

## Author Contributions

JCC – Conceptualization, Data curation, Formal Analysis, Methodology, Supervision, Visualization, Writing – original draft, Writing – review & editing

MK – Methodology, Software, Writing – review & editing

SL – Methodology, Software, Writing – review & editing

KA – Formal Analysis, Writing – review & editing

JZZ – Methodology, Software

LMS – Visualization, Writing – review & editing

TJY – Visualization, Writing – review & editing

EAC – Formal Analysis

DAD – Formal Analysis

HC – Data curation, Resources, Investigation

TN – Project administration

MSR – Data curation, Project administration

SV – Project administration, Writing – review & editing

RD – Data curation, Resources, Investigation, Funding acquisition, Writing – review & editing

NLH – Data curation, Funding acquisition, Project administration, Writing – review & editing

STM – Data curation, Funding acquisition, Project administration, Writing – review & editing

DEW – Conceptualization, Funding acquisition, Methodology, Project administration, Supervision, Writing – review & editing

RLM – Conceptualization, Funding acquisition, Methodology, Project administration, Supervision, Writing – review & editing

## Declaration of Interests

The authors declare no competing interests.

## Data and code availability

Upon publication, the Minimac files for the *Soifua Manuia* reference panel will be made available through dbGaP (accession number: phs000972.v5.p1). The discovery cohort data used for this study are available through dbGaP (accession numbers: phs000914.v1.p1 and phs000972.v5.p1). A template for the imputation code is available at https://github.com/jennaccarlson/imputation; additional code for creating the imputation reference panels and imputation quality analysis will be made available upon request.

## Notes

### Competing Interest Statement

The authors have declared no competing interest.

### Author Declarations

This study has been approved by the Health Research Committee of the Samoa Ministry of Health and the institutional review boards of Brown University and the University of Pittsburgh. All participants gave informed consent.

### Summary of Updates

New analyses performed: genome-wide comparisons, genotype concordance, enrichment of variants, phasing comparison, and meta imputation

## References

1. 1000 Genomes Project Consortium, Auton, A., Brooks, L.D., Durbin, R.M., Garrison, E.P., Kang, H.M., Korbel, J.O., Marchini, J.L., McCarthy, S., McVean, G.A., et al. (2015). A global reference for human genetic variation. Nature 526, 68–74. 10.1038/nature15393.

2. McCarthy, S., Das, S., Kretzschmar, W., Delaneau, O., Wood, A.R., Teumer, A., Kang, H.M., Fuchsberger, C., Danecek, P., Sharp, K., et al. (2016). A reference panel of 64,976 haplotypes for genotype imputation. Nature Genetics 48, 1279–1283. 10.1038/ng.3643.

3. Taliun, D., Harris, D.N., Kessler, M.D., Carlson, J., Szpiech, Z.A., Torres, R., Taliun, S.A.G., Corvelo, A., Gogarten, S.M., Kang, H.M., et al. (2021). Sequencing of 53,831 diverse genomes from the NHLBI TOPMed Program. Nature 590, 290–299. 10.1038/s41586-021-03205-y.

4. Kowalski, M.H., Qian, H., Hou, Z., Rosen, J.D., Tapia, A.L., Shan, Y., Jain, D., Argos, M., Arnett, D.K., Avery, C., et al. (2019). Use of >100,000 NHLBI Trans-Omics for Precision Medicine (TOPMed) Consortium whole genome sequences improves imputation quality and detection of rare variant associations in admixed African and Hispanic/Latino populations. PLoS Genet 15, e1008500. 10.1371/journal.pgen.1008500.

5. Friedlaender, J.S., Friedlaender, F.R., Reed, F.A., Kidd, K.K., Kidd, J.R., Chambers, G.K., Lea, R.A., Loo, J.-H., Koki, G., Hodgson, J.A., et al. (2008). The Genetic Structure of Pacific Islanders. PLOS Genetics 4, e19. 10.1371/journal.pgen.0040019.

6. Skoglund, P., Posth, C., Sirak, K., Spriggs, M., Valentin, F., Bedford, S., Clark, G.R., Reepmeyer, C., Petchey, F., Fernandes, D., et al. (2016). Genomic insights into the peopling of the Southwest Pacific. Nature 538, 510–513. 10.1038/nature19844.

7. Tsai, H.-J., Sun, G., Smelser, D., Viali, S., Tufa, J., Jin, L., Weeks, D.E., McGarvey, S.T., and Deka, R. (2004). Distribution of genome-wide linkage disequilibrium based on microsatellite loci in the Samoan population. Human Genomics 1, 327. 10.1186/1479-7364-1-5-327.

8. Harris, D.N., Kessler, M.D., Shetty, A.C., Weeks, D.E., Minster, R.L., Browning, S., Cochrane, E.E., Deka, R., Hawley, N.L., Reupena, M.S., et al. (2020). Evolutionary history of modern Samoans. Proceedings of the National Academy of Sciences 117, 201913157. 10.1073/pnas.1913157117.

9. World Health Organization. Regional Office for the Western Pacific (2021). Progress on the prevention and control of noncommunicable diseases in the Western Pacific Region: country capacity survey 2019 (WHO Regional Office for the Western Pacific).

10. Graham, G. (2015). Disparities in Cardiovascular Disease Risk in the United States. Current Cardiology Reviews 11, 238–245.

11. Anand, S., Bradshaw, C., and Prabhakaran, D. (2020). Prevention and management of CVD in LMICs: why do ethnicity, culture, and context matter? BMC Med 18, 7. 10.1186/s12916-019-1480-9.

12. Hawley, N.L., Minster, R.L., Weeks, D.E., Viali, S., Reupena, M.S., Sun, G., Cheng, H., Deka, R., and Mcgarvey, S.T. (2014). Prevalence of adiposity and associated cardiometabolic risk factors in the samoan genome-wide association study. American Journal of Human Biology 26, 491–501. 10.1002/ajhb.22553.

13. McGarvey, S.T. (2001). Cardiovascular disease (CVD) risk factors in Samoa and American Samoa, 1990-95. Pacific health dialog 8, 157–162.

14. Klag, M.J., Whelton, P.K., Randall, B.L., Neaton, J.D., Brancati, F.L., and Stamler, J. (1997). End-stage Renal Disease in African-American and White Men: 16-Year MRFIT Findings. JAMA 277, 1293–1298. 10.1001/jama.1997.03540400043029.

15. SIGMA Type 2 Diabetes Consortium, Williams, A.L., Jacobs, S.B., Moreno-Macías, H., Huerta-Chagoya, A., Churchhouse, C., Márquez-Luna, C., García-Ortíz, H., Gómez-Vázquez, M.J., Burtt, N.P., et al. (2014). Sequence variants in SLC16A11 are a common risk factor for type 2 diabetes in Mexico. Nature 506, 97–101. 10.1038/nature12828.

16. Nair, A.K., and Baier, L.J. (2015). Complex Genetics of Type 2 Diabetes and Effect Size: What have We Learned from Isolated Populations? Rev Diabet Stud 12, 299–319. 10.1900/RDS.2015.12.299.

17. Staimez, L.R., Deepa, M., Ali, M.K., Mohan, V., Hanson, R.L., and Narayan, K.M.V. (2019). Tale of two Indians: Heterogeneity in type 2 diabetes pathophysiology. Diabetes/Metabolism Research and Reviews 35, e3192. 10.1002/dmrr.3192.

18. Vartiainen, E., Dianjun, D., Marks, J.S., Korhonen, H., Guanyi, G., Ze-Yu, G., Koplan, J.P., Pietinen, P., Guang-Lin, W., Williamson, D., et al. (1991). Mortality, cardiovascular risk factors, and diet in China, Finland, and the United States. Public Health Rep 106, 41–46.

19. Gupta, A.K., Poulter, N.R., Dobson, J., Eldridge, S., Cappuccio, F.P., Caulfield, M., Collier, D., Cruickshank, J.K., Sever, P.S., Feder, G., et al. (2010). Ethnic Differences in Blood Pressure Response to First and Second-Line Antihypertensive Therapies in Patients Randomized in the ASCOT Trial. American Journal of Hypertension 23, 1023–1030. 10.1038/ajh.2010.105.

20. Ojji, D.B., Mayosi, B., Francis, V., Badri, M., Cornelius, V., Smythe, W., Kramer, N., Barasa, F., Damasceno, A., Dzudie, A., et al. (2019). Comparison of Dual Therapies for Lowering Blood Pressure in Black Africans. New England Journal of Medicine 380, 2429–2439. 10.1056/NEJMoa1901113.

21. James, P.A., Oparil, S., Carter, B.L., Cushman, W.C., Dennison-Himmelfarb, C., Handler, J., Lackland, D.T., LeFevre, M.L., MacKenzie, T.D., Ogedegbe, O., et al. (2014). 2014 Evidence-Based Guideline for the Management of High Blood Pressure in Adults: Report From the Panel Members Appointed to the Eighth Joint National Committee (JNC 8). JAMA 311, 507–520. 10.1001/jama.2013.284427.

22. Mills, M.C., and Rahal, C. (2020). The GWAS Diversity Monitor tracks diversity by disease in real time. Nat Genet 52, 242–243. 10.1038/s41588-020-0580-y.

23. Precision medicine needs an equity agenda (2021). Nat Med 27, 737–737. 10.1038/s41591-021-01373-y.

24. Burke, W. (2021). Utility and Diversity: Challenges for Genomic Medicine. Annual Review of Genomics and Human Genetics 22, 1–24. 10.1146/annurev-genom-120220-082640.

25. McGarvey, S.T., Levinson, P.D., Bausser-Man, L., Galanis, D.J., and Hornick, C.A. (1993). Population change in adult obesity and blood lipids in American Samoa from 1976–1978 to 1990. American Journal of Human Biology 5, 17–30. 10.1002/ajhb.1310050106.

26. Chin-Hong, P.V., and McGarvey, S.T. (1996). Lifestyle incongruity and adult blood pressure in Western Samoa. Psychosomatic medicine 58, 131–137.

27. Galanis, D.J., McGarvey, S.T., Quested, C., Sio, B., and Afele-Fa’amuli, S.A. (1999). Dietary intake of modernizing Samoans: implications for risk of cardiovascular disease. Journal of the American Dietetic Association 99, 184–190.

28. Dai, F., Keighley, E.D., Sun, G., Indugula, S.R., Roberts, S.T., Åberg, K., Smelser, D., Tuitele, J., Jin, L., Deka, R., et al. (2007). Genome-wide scan for adiposity-related phenotypes in adults from American Samoa. International Journal of Obesity 31, 1832– 1842. 10.1038/sj.ijo.0803675.

29. Dai, F., Sun, G., Åberg, K., Keighley, E.D., Indugula, S.R., Roberts, S.T., Smelser, D., Viali, S., Jin, L., Deka, R., et al. (2008). A Whole Genome Linkage Scan Identifies Multiple Chromosomal Regions Influencing Adiposity-Related Traits among Samoans. Annals of Human Genetics 72, 780–792. 10.1111/j.1469-1809.2008.00462.x.

30. Minster, R.L., Hawley, N.L., Su, C.-T., Sun, G., Kershaw, E.E., Cheng, H., Buhule, O.D., Lin, J., Reupena, M.S., Viali, S., et al. (2016). A thrifty variant in CREBRF strongly influences body mass index in Samoans. Nature Genetics 48, 1049–1054. 10.1038/ng.3620.

31. Carlson, J.C., Krishnan, M., Rosenthal, S.L., Russell, E.M., Zhang, J.Z., Hawley, N.L., Moors, J., Cheng, H., Dalbeth, N., de Zoysa, J.R., et al. (2023). A stop-gain variant in *BTNL9* is associated with atherogenic lipid profiles. HGG Adv 4, 100155. 10.1016/j.xhgg.2022.100155.

32. Laurie, C.C., Doheny, K.F., Mirel, D.B., Pugh, E.W., Bierut, L.J., Bhangale, T., Boehm, F., Caporaso, N.E., Cornelis, M.C., Edenberg, H.J., et al. (2010). Quality control and quality assurance in genotypic data for genome-wide association studies. Genetic Epidemiology 34, 591–602. 10.1002/gepi.20516.

33. Gusev, A., Shah, M.J., Kenny, E.E., Ramachandran, A., Lowe, J.K., Salit, J., Lee, C.C., Levandowsky, E.C., Weaver, T.N., Doan, Q.C., et al. (2012). Low-Pass Genome-Wide Sequencing and Variant Inference Using Identity-by-Descent in an Isolated Human Population. Genetics 190, 679–689. 10.1534/genetics.111.134874.

34. Loh, P.-R., Danecek, P., Palamara, P.F., Fuchsberger, C., A Reshef, Y., K Finucane, H., Schoenherr, S., Forer, L., McCarthy, S., Abecasis, G.R., et al. (2016). Reference-based phasing using the Haplotype Reference Consortium panel. Nat Genet 48, 1443–1448. 10.1038/ng.3679.

35. Deelen, P., Bonder, M.J., Van Der Velde, K.J., Westra, H.J., Winder, E., Hendriksen, D., Franke, L., and Swertz, M.A. (2014). Genotype harmonizer: Automatic strand alignment and format conversion for genotype data integration. BMC Research Notes 7, 901. 10.1186/1756-0500-7-901.

36. Das, S., Forer, L., Schönherr, S., Sidore, C., Locke, A.E., Kwong, A., Vrieze, S.I., Chew, E.Y., Levy, S., McGue, M., et al. (2016). Next-generation genotype imputation service and methods. Nature Genetics 48, 1284–1287. 10.1038/ng.3656.

37. Yu, K., Das, S., LeFaive, J., Kwong, A., Pleiness, J., Forer, L., Schönherr, S., Fuchsberger, C., Smith, A.V., and Abecasis, G.R. (2022). Meta-imputation: An efficient method to combine genotype data after imputation with multiple reference panels. The American Journal of Human Genetics 109, 1007–1015. 10.1016/j.ajhg.2022.04.002.

38. McLaren, W., Gil, L., Hunt, S.E., Riat, H.S., Ritchie, G.R.S., Thormann, A., Flicek, P., and Cunningham, F. (2016). The Ensembl Variant Effect Predictor. Genome Biology 17, 122. 10.1186/s13059-016-0974-4.

39. Cingolani, P., Platts, A., Wang, L.L., Coon, M., Nguyen, T., Wang, L., Land, S.J., Lu, X., and Ruden, D.M. (2012). A program for annotating and predicting the effects of single nucleotide polymorphisms, SnpEff. Fly (Austin) 6, 80–92. 10.4161/fly.19695.

40. Danecek, P., Auton, A., Abecasis, G., Albers, C.A., Banks, E., DePristo, M.A., Handsaker, R.E., Lunter, G., Marth, G.T., Sherry, S.T., et al. (2011). The variant call format and VCFtools. Bioinformatics 27, 2156–2158. 10.1093/bioinformatics/btr330.

41. Yang, J., Lee, S.H., Goddard, M.E., and Visscher, P.M. (2011). GCTA: A Tool for Genome-wide Complex Trait Analysis. Am J Hum Genet 88, 76–82. 10.1016/j.ajhg.2010.11.011.

42. Danecek, P., Bonfield, J.K., Liddle, J., Marshall, J., Ohan, V., Pollard, M.O., Whitwham, A., Keane, T., McCarthy, S.A., Davies, R.M., et al. (2021). Twelve years of SAMtools and BCFtools. Gigascience 10, giab008. 10.1093/gigascience/giab008.

43. The NHLBI Trans-Omics for Precision Medicine (TOPMed) Whole Genome Sequencing Program. (2018). BRAVO variant browser: University of Michigan and NHLBI.

44. Yang, M.-Y., Zhong, J.-D., Li, X., Tian, G., Bai, W.-Y., Fang, Y.-H., Qiu, M.-C., Yuan, C.-D., Yu, C.-F., Li, N., et al. (2024). SEAD reference panel with 22,134 haplotypes boosts rare variant imputation and genome-wide association analysis in Asian populations. Nat Commun 15, 10839. 10.1038/s41467-024-55147-4.

