## Supplement for "The *Soifua Manuia* reference panel with 2,570 Samoan haplotypes improves genotype imputation quality among Samoans"

### Supplemental Information

#### Table of Contents

Abstract Translated to Samoan

Supplemental Figures

- Figure S1. Principal components of ancestry for participants.
- Figure S2. Information content for participants
- Figure S3. Median imputation quality versus minor allele frequency by reference panel.
- Figure S4. Median empirical r2 versus minor allele frequency by reference panel for genotyped variants.
- Figure S5. Median imputation quality versus fixation index by reference panel.
- Figure S6. Median imputation quality versus fixation index by reference panel and MAF bin.
- Figure S7. Median imputation quality versus LD score by reference panel.
- Figure S8. Median imputation quality versus LD score by reference panel and MAF bin.
- Figure S9. Imputation quality versus minor allele frequency by reference panel.
- Figure S10. Imputation quality versus minor allele frequency for meta-imputation.
- Figure S11. Count of well imputed variants from meta imputation.
- Figure S12. Imputation quality versus minor allele frequency for meta-imputation.

Supplemental Tables

- Table S1. Number of well-imputed variants by reference panel.
- Table S2. Number of rescued variants by reference panel.
- Table S3. Consequence and variant groupings used to test enrichment of variants.
- Table S4. Enrichment in variant consequence and impact for Samoan-specific imputation.
- Table S5. Enrichment in variant consequence and impact for Samoan-specific imputation.
- Table S6. Mean $r^2$ by reference panel.
- Table S7. Median $r^2$ by reference panel.
- Table S8. Median $F_{ST}$ and LD score by minor allele frequency (MAF) and chromosome.
- Table S9. Number of well-imputed variants by reference panel for meta imputation.

#### Abstract Translated to Samoan

[Will be translated prior to publication, upon acceptance of the manuscript]

Genotype imputation is fundamental to association studies, and yet even gold standard panels like TOPMed are limited in the populations for which they yield good imputation. Specifically, Pacific Islanders are poorly represented in extant panels. To address this, we constructed an imputation reference panel using 1,285 Samoan individuals with whole-genome sequencing, combined with 1000 Genomes (1KGP) individuals, to create a reference panel that better represents Pacific Islander, specifically Samoan, genetic variation. We compared this panel to 1KGP and TOPMed-R3 panels based on imputed variants using genotyping array data for 1,834 Samoan participants who were not part of the panels. The 1KGP + 1285 Samoan panel yielded up to two times more well-imputed ($r^2 \geq 0.80$) variants than TOPMed-R3 and 1KGP and was enriched for moderate and high impact variants. There was improved imputation accuracy across the minor allele frequency (MAF) spectrum, although it was more pronounced for variants with $0.01 \leq MAF \leq 0.05$. Imputation accuracy ($r^2$) was greater for population-specific variants (high fixation index, $F_{ST}$) and those from larger haplotypes (high LD score). However, the gain in imputation accuracy over TOPMed-R3 was largest for small haplotypes (low LD score), reflecting the Samoan panel's ability to capture population-specific variation not well tagged by other panels. We also augmented the 1KGP reference panel with varying numbers of Samoan participants and found that panels with 24 Samoans yielded similar performance to TOPMed-R3, and panels with 48 or more Samoans included outperformed TOPMed-R3 for all variants with $MAF \geq 0.001$. Meta imputation of the TOPMed-R3 and 1285 Samoan panels yielded poorer performance than the Samoan only panel. We also demonstrated that the phasing of the reference panel impacts the imputation of population-specific variants when the reference panel is composed of individuals from an isolated population and not combined with ancestrally diverse haplotypes. This study identifies variants with improved imputation using population-specific reference panels and provides a framework for constructing other population-specific reference panels.

### Supplemental Figures

**Figure S1. Principal components of ancestry for participants.** The first two principal components of ancestry are plotted. Blue points indicate participants with WGS (n=1,285) and the gray points indicate those with genotyping array data only (n=1,834).

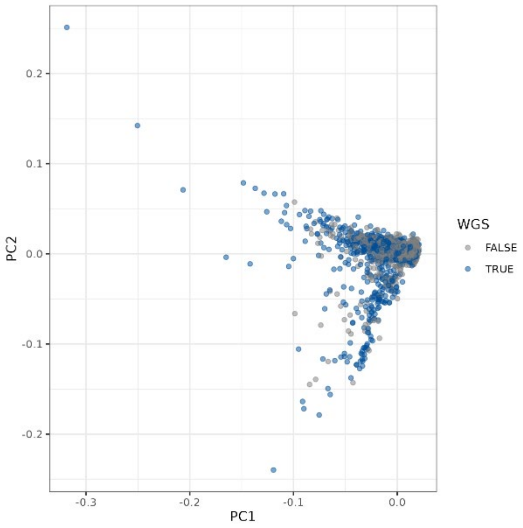


**Figure S2. Information content for participants.** The subset of 1,285 participants to send for whole-genome sequencing were chosen through INFOSTIP rankings. The total information content obtained from INFOSTIP is plotted cumulatively over the 1,285 participants sent for whole-genome sequencing.

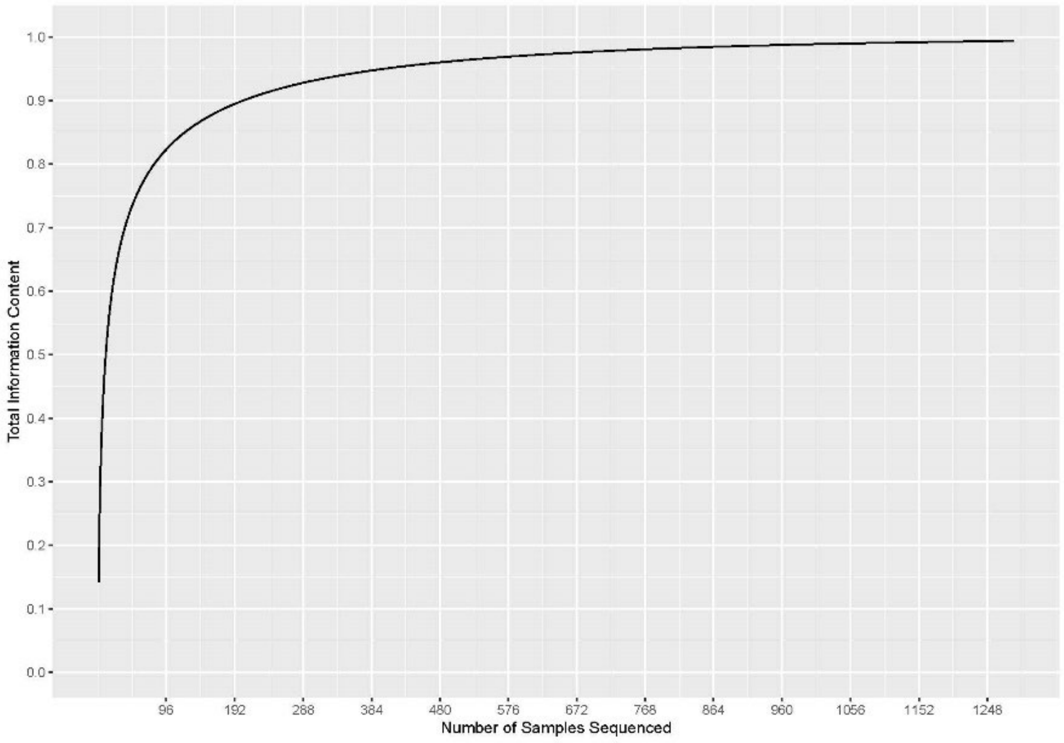

**Figure S3. Median imputation quality versus minor allele frequency by reference panel.** Median imputation quality ($r^2$) by minor allele frequency (MAF) bin (left column: based on 1KGP + 1285S imputation; right column: based on the TOPMed imputation) is plotted by reference panel on chromosomes 5 and 21.

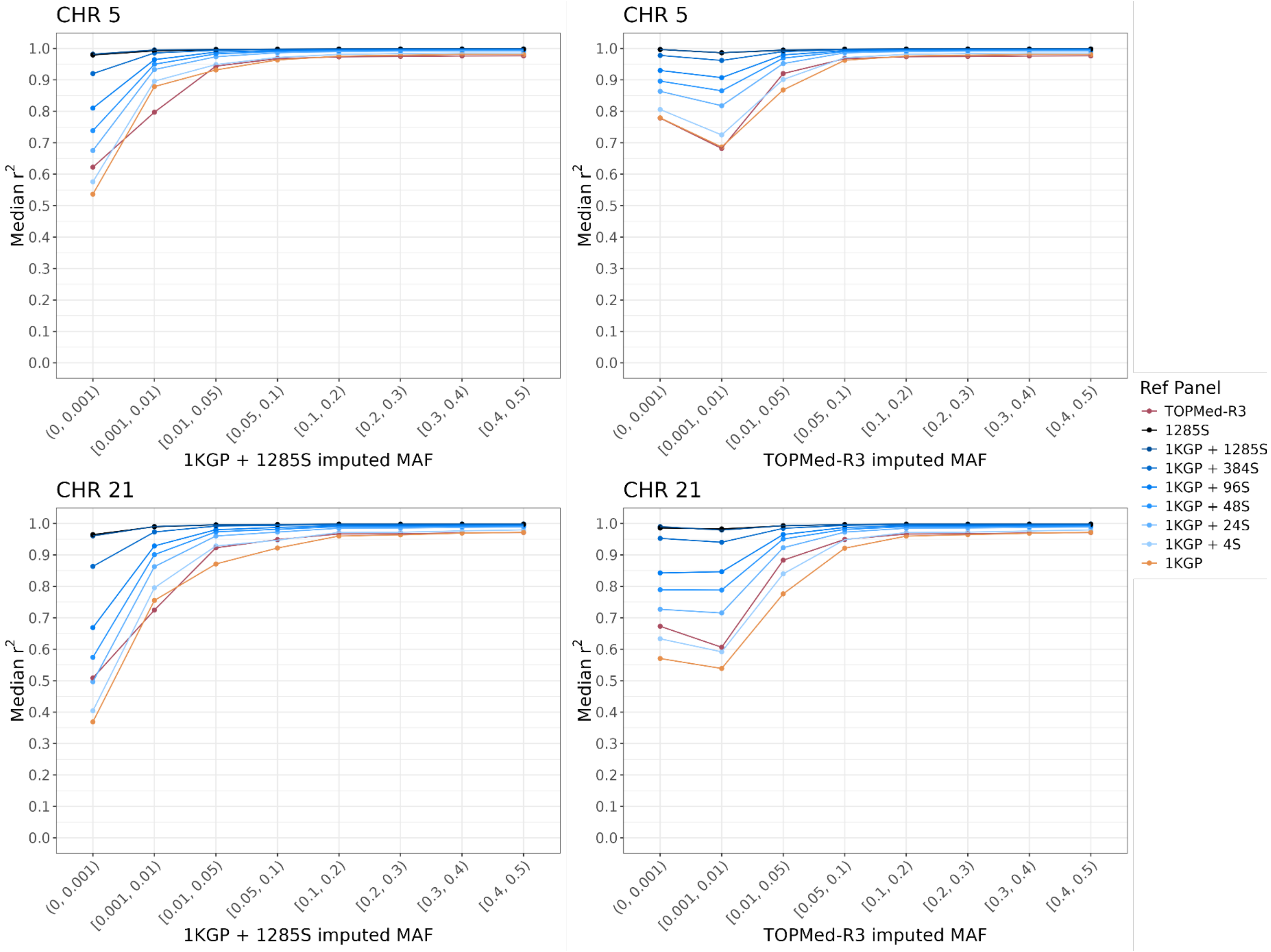

**Figure S4. Median empirical r2 versus minor allele frequency by reference panel for genotyped variants.** Median empirical $r^2$ by minor allele frequency (MAF) bin (based on 1KGP + 1285S imputation) is plotted by reference panel on chromosomes 5 and 21. Y-axis limited to 0.8-1.0 to show differences across panels.

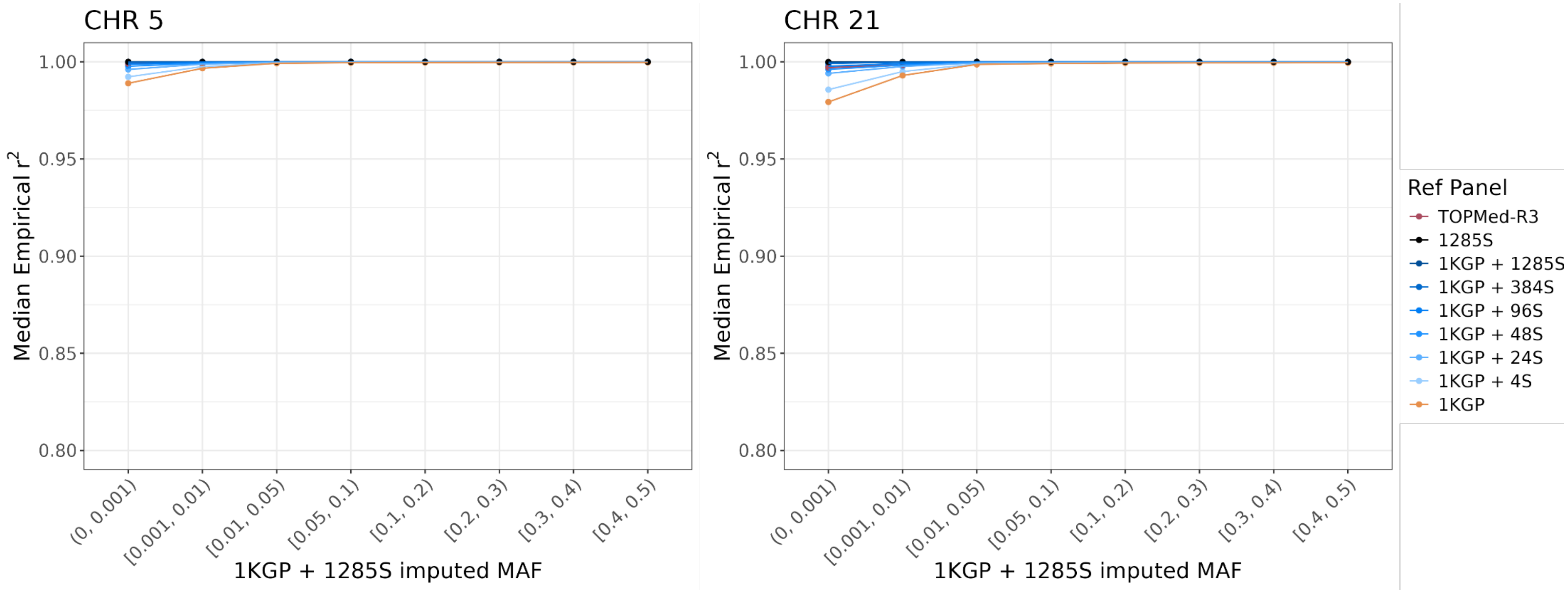

**Figure S5. Median imputation quality versus fixation index by reference panel.**

Median imputation quality ($r^2$) by empirical Weir and Cockerham Fixation Index ($F_{ST}$) deciles is plotted by reference panel on chromosomes 5 and 21.

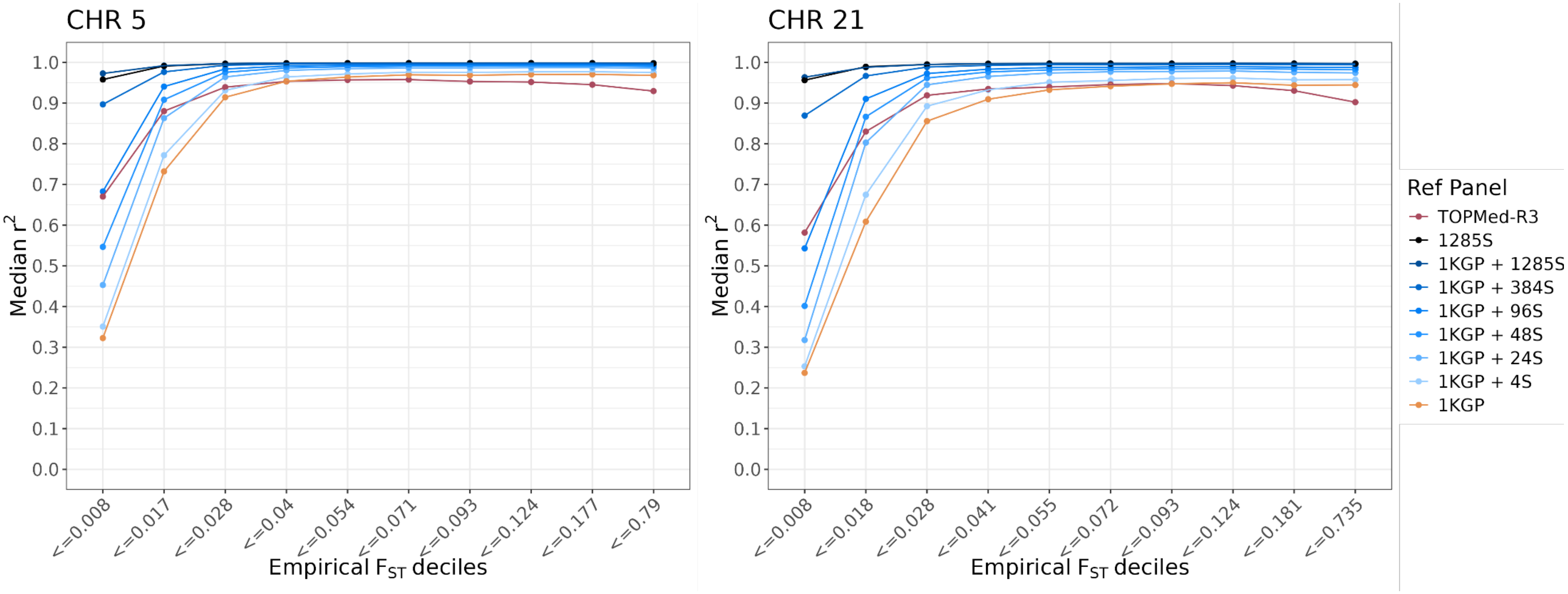

**Figure S6. Median imputation quality versus fixation index by reference panel and MAF bin.** Median imputation quality (r2) by median-stratified Weir and Cockerham Fixation Index (FST) across MAF (based on 1KGP + 1285S imputation) is plotted by reference panel on chromosomes 5 and 21. FST was stratified into 'low' and 'high' about the median FST within each MAF bin (values given in Table S5).

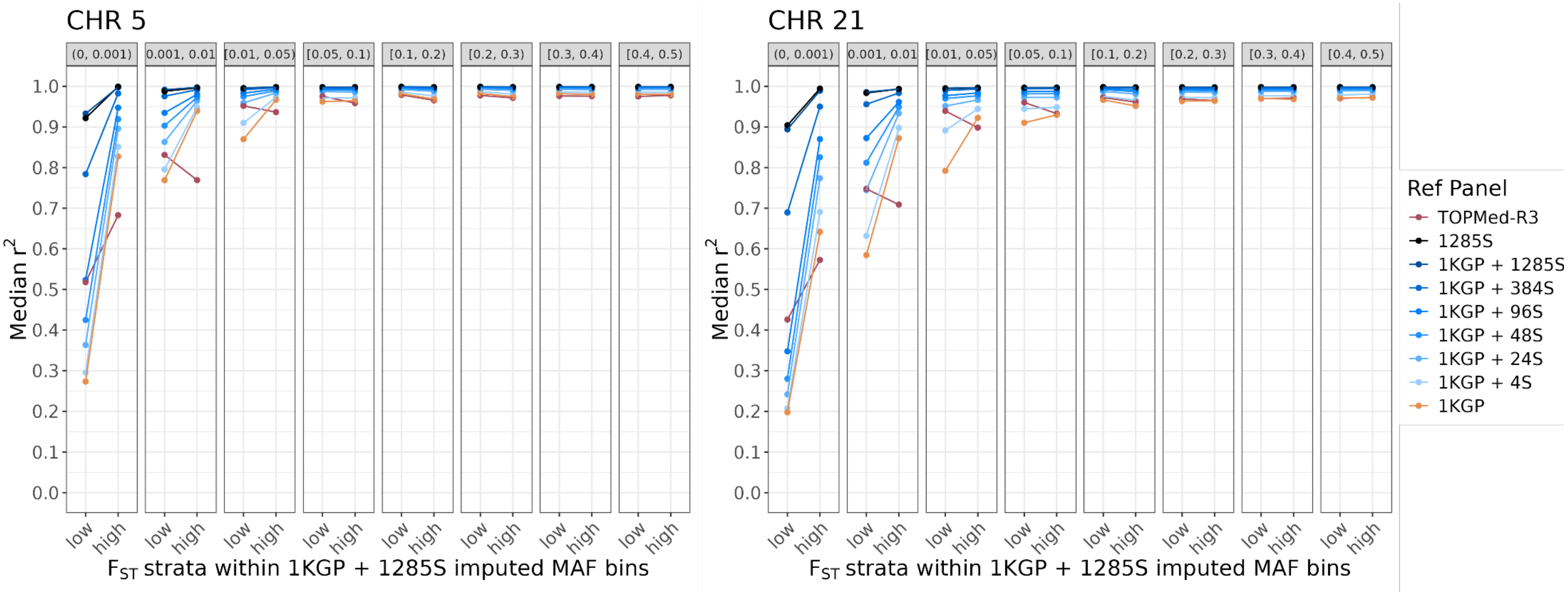

**Figure S7. Median imputation quality versus LD score by reference panel.** Median imputation quality ($r^2$) by empirical LD score deciles is plotted by reference panel on chromosomes 5 and 21.

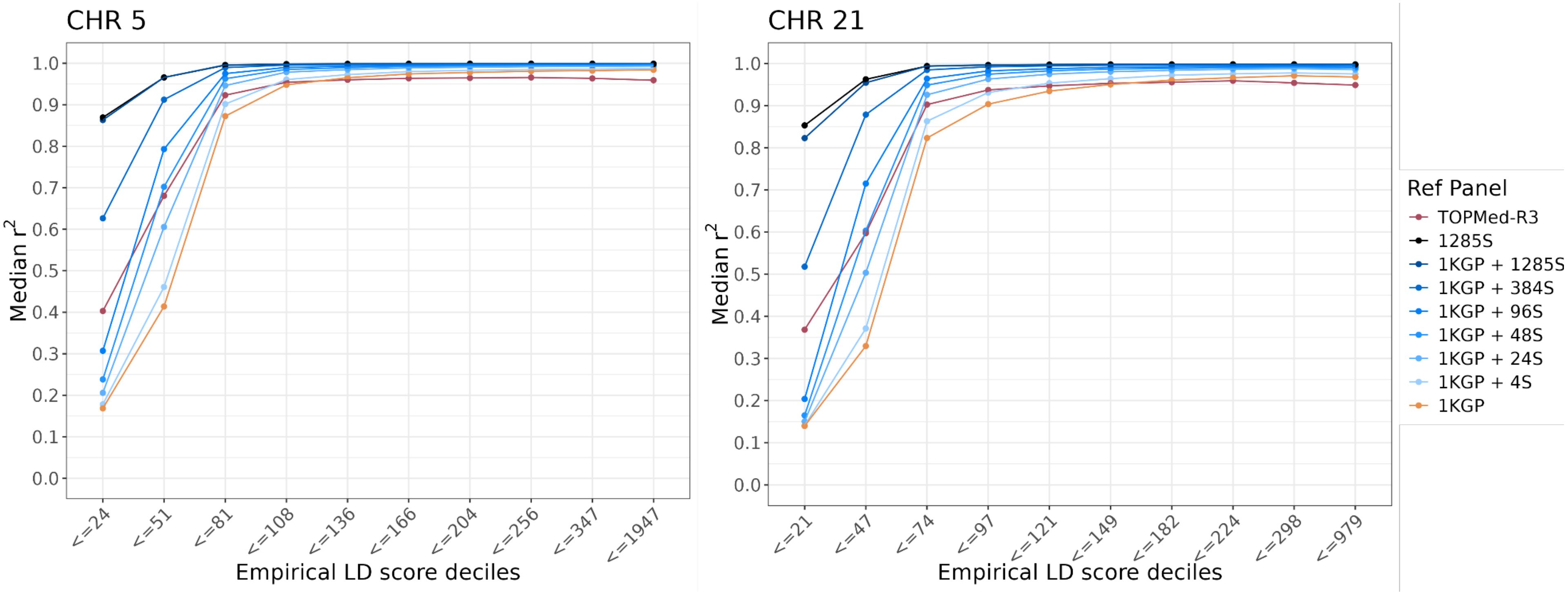

**Figure S8. Median imputation quality versus LD score by reference panel and MAF bin.** Mean imputation quality ($r^2$) by median-stratified LD score across MAF (based on 1KGP + 1285S imputation) is plotted by reference panel on chromosomes 5 and 21. LD score was stratified into 'low' and 'high' about the median LD score within each MAF bin (values given in Table S5).

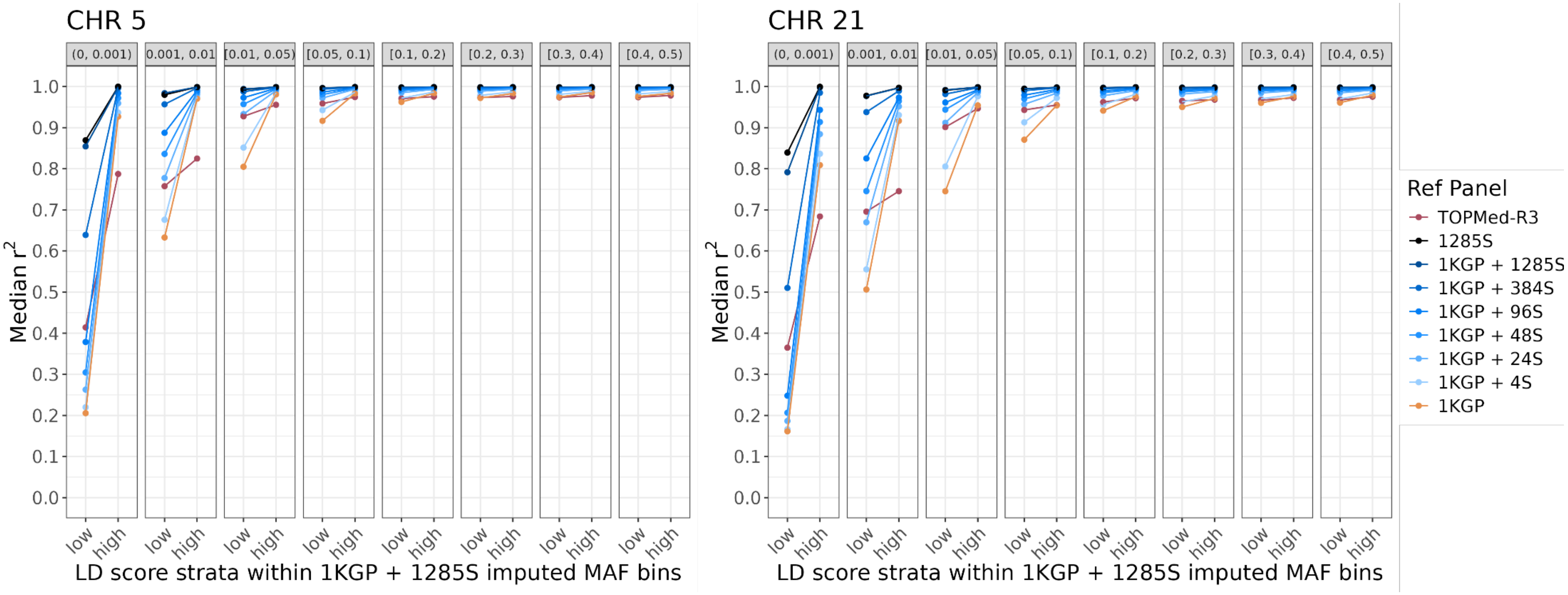

**Figure S9. Imputation quality versus minor allele frequency by reference panel.** Mean imputation quality ($r^2$) by minor allele frequency (MAF) bin (based on 1KGP + 1285S imputation) is plotted by reference panel across chromosomes 1-22 for a subset of n=100 Samoans.

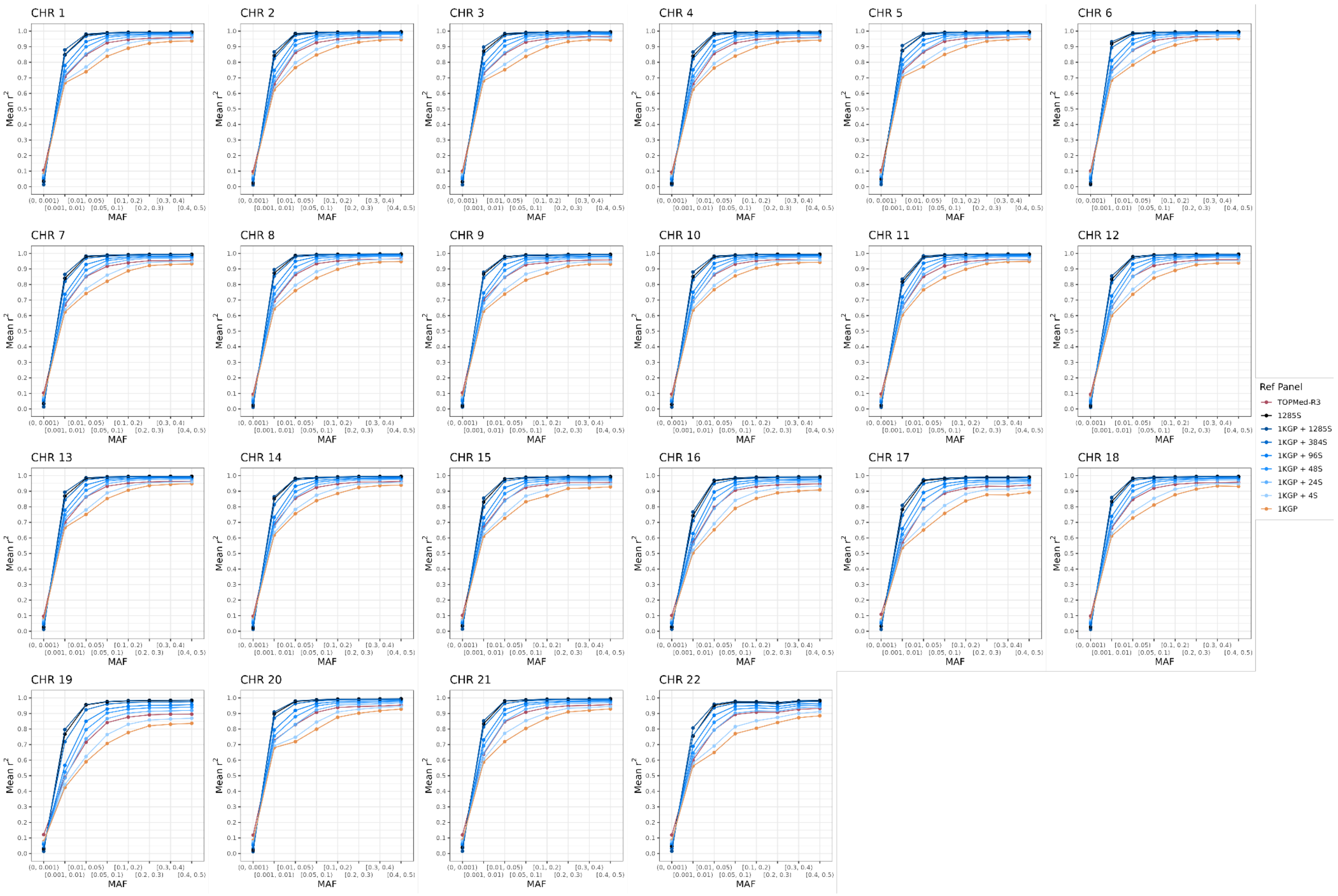

**Figure S10. Imputation quality versus minor allele frequency for meta-imputation.** Mean imputation quality ($r^2$) by minor allele frequency (MAF) bin (based on 1285S imputation) is plotted for meta-imputation (purple) of TOPMed-R3 (red) and 1285S (black) reference panels for chromosomes 5 and 21 for *n*=1,834 Samoans.

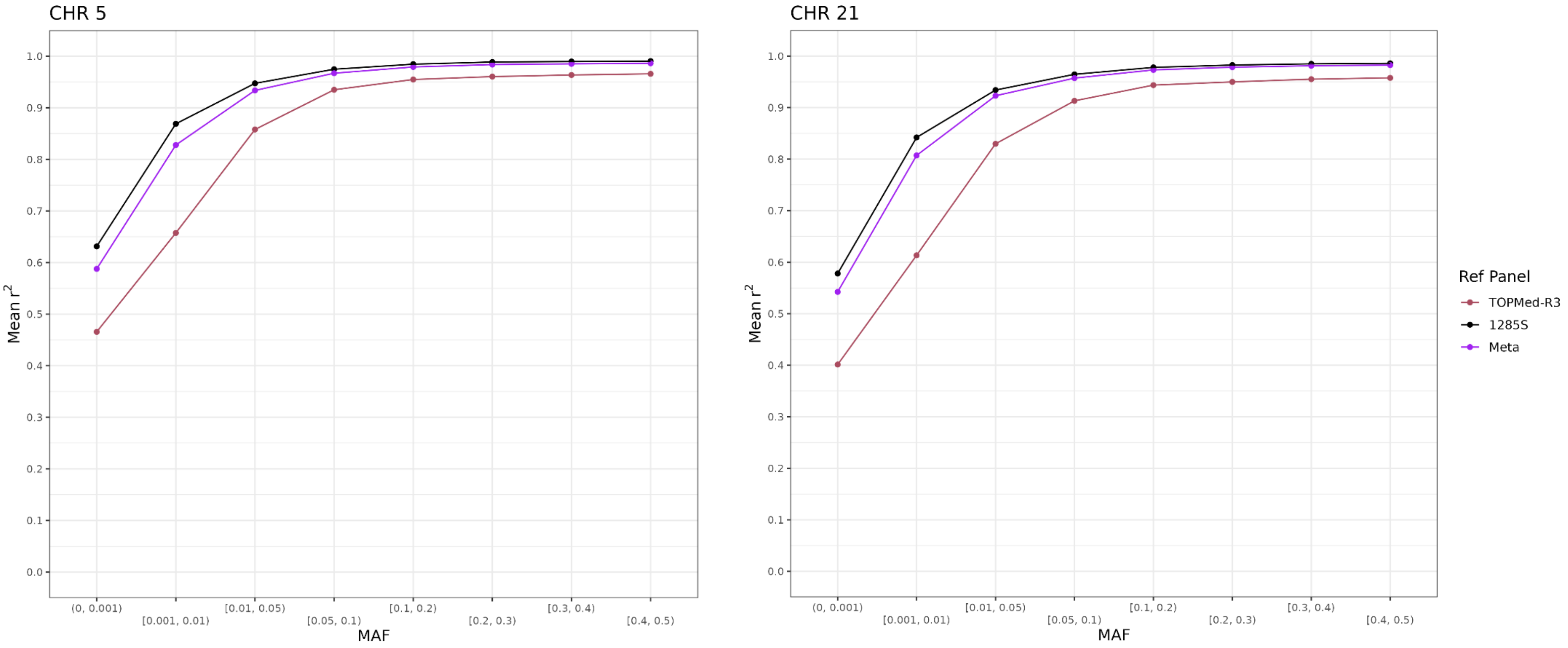

**Figure S11. Count of well imputed variants from meta imputation.** The number of variants with imputation quality $r^2 \geq 0.80$ from TOPMed-R3, 1285S and meta imputation is plotted across minor allele frequency (MAF) bins for chromosomes 5 and 21.

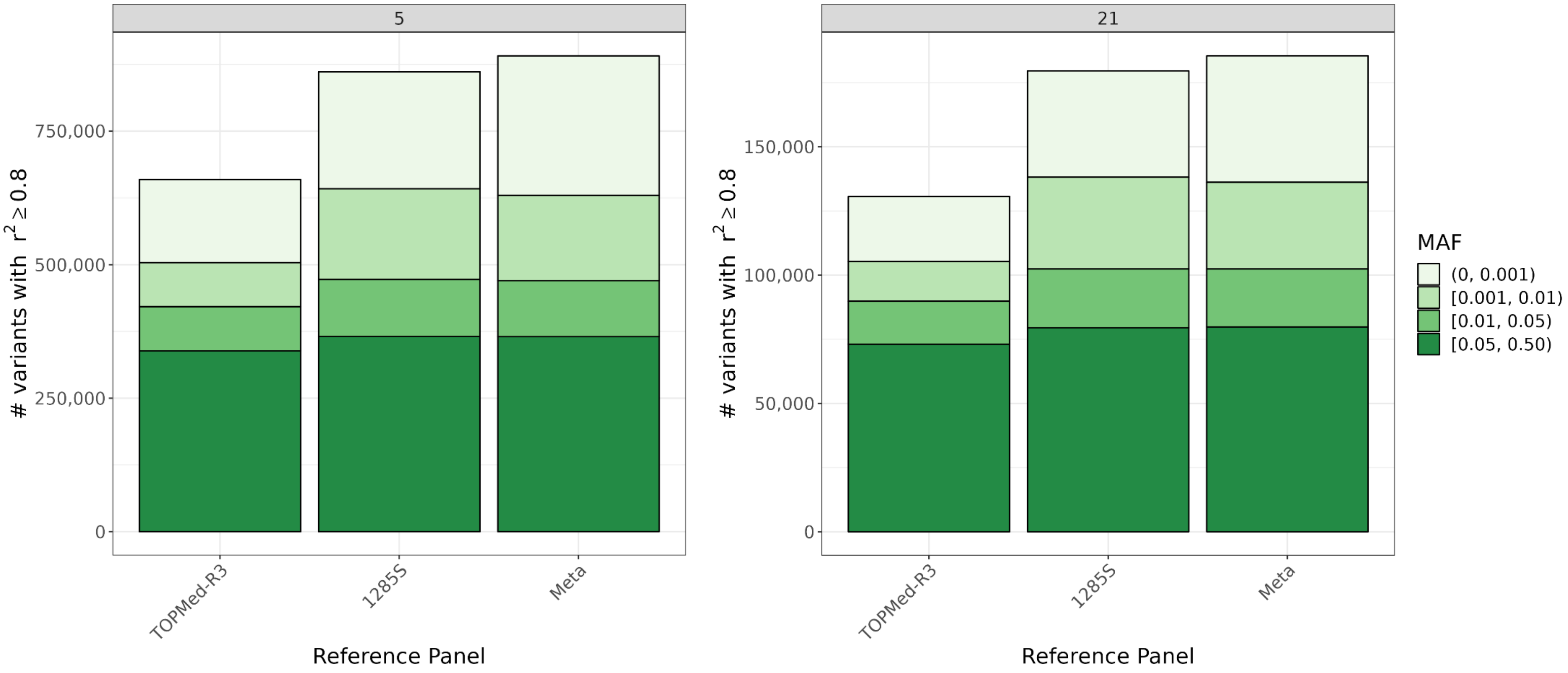

**Figure S12. Imputation quality versus minor allele frequency for meta-imputation.** Mean imputation quality ($r^2$) by minor allele frequency (MAF) bin (based on 1285S imputation) is plotted for meta-imputation (purple) of TOPMed-R3 (red) and 1285S (black) reference panels across chromosomes 1-22 for a subset of n=100 Samoans.

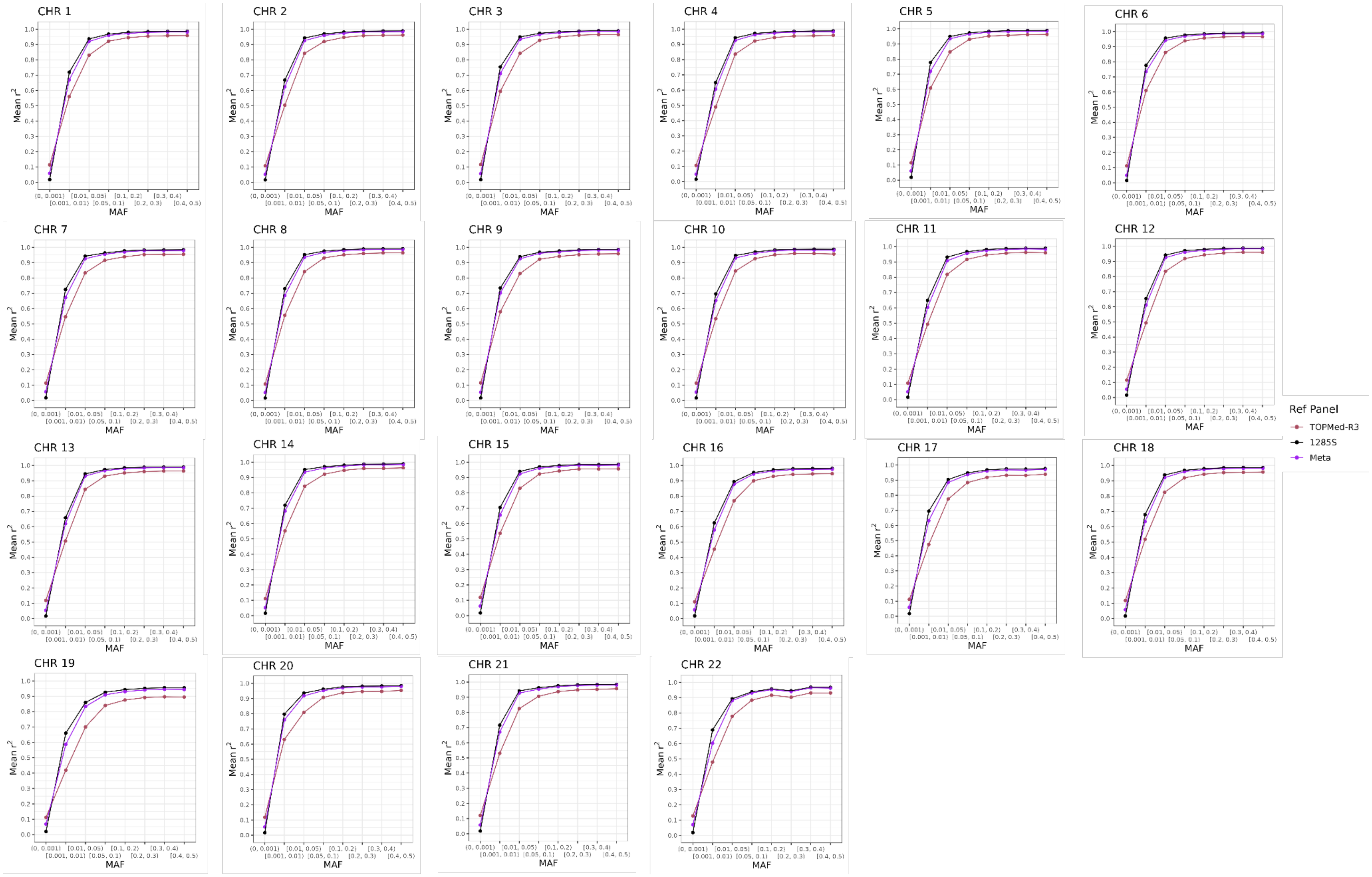

#### Supplemental Tables

**Table S1. Number of well-imputed variants by reference panel.** Number of well-imputed variants ($r^2 \geq 0.80$) by minor allele frequency (MAF) across reference panels and chromosome.

| **CHR 5** | | | **MAF bin** | | |
|---|---|---|---|---|---|
| **Reference Panel** | **(0, 0.001)** | **[0.001, 0.01)** | **[0.01, 0.05)** | **[0.05, 0.50)** | **Total** |
| TOPMed-R3 | 155,605 | 82,367 | 82,544 | 338,837 | 659,353 |
| 1KGP | 131,625 | 79,984 | 55,619 | 317,630 | 584,858 |
| 1KGP + 4S | 136,626 | 84,602 | 63,674 | 334,121 | 619,023 |
| 1KGP + 24S | 152,224 | 100,462 | 86,522 | 357,938 | 697,146 |
| 1KGP + 24S | 164,942 | 115,457 | 98,489 | 363,249 | 742,137 |
| 1KGP + 96S | 184,106 | 134,454 | 106,917 | 365,778 | 791,255 |
| 1KGP + 384S | 244,995 | 181,799 | 112,739 | 367,185 | 906,718 |
| 1KGP + 1285S | 274,721 | 202,490 | 113,038 | 365,562 | 955,811 |
| 1KGP + 1285S in-house phasing | 274,889 | 202,225 | 113,013 | 365,561 | 955,688 |
| 1285S | 268,008 | 205,252 | 113,700 | 367,448 | 954,408 |
| 1285S in-house phasing | 193,663 | 188,861 | 112,359 | 365,440 | 860,323 |
| **CHR 21** | | | **MAF bin** | | |
| **Reference Panel** | **(0, 0.001)** | **[0.001, 0.01)** | **[0.01, 0.05)** | **[0.05, 0.50)** | **Total** |
| TOPMed-R3 | 25,364 | 15,436 | 16,752 | 73,091 | 130,643 |
| 1KGP | 20,867 | 14,400 | 10,779 | 65,506 | 111,552 |
| 1KGP + 4S | 22,233 | 15,816 | 13,837 | 70,073 | 121,959 |
| 1KGP + 24S | 24,890 | 19,448 | 18,819 | 76,875 | 140,032 |
| 1KGP + 24S | 27,382 | 23,166 | 21,190 | 78,562 | 150,300 |
| 1KGP + 96S | 31,474 | 27,052 | 22,647 | 79,410 | 160,583 |
| 1KGP + 384S | 46,338 | 38,460 | 24,355 | 79,998 | 189,151 |
| 1KGP + 1285S | 52,798 | 43,451 | 24,488 | 79,652 | 200,389 |
| 1KGP + 1285S in-house phasing | 52,802 | 43,473 | 24,492 | 79,651 | 200,418 |
| 1285S | 53,581 | 44,348 | 24,743 | 80,248 | 202,920 |
| 1285S in-house phasing | 37,376 | 39,500 | 24,090 | 79,576 | 180,542 |

**Table S2. Number of rescued variants by reference panel.** Number and percent of imputed variants that were low-quality ($r^2 < 0.80$) in the 1KGP imputation but were well-imputed ($r^2 \geq 0.80$) by minor allele frequency (MAF) across reference panels and chromosome.

| **CHR 5** | **Reference Panel** | | | | | | | | | |
|---|---|---|---|---|---|---|---|---|---|---|
| **MAF** ***# variants lost in 1KGP*** | **1KGP + 4S** | **1KGP + 24S** | **1KGP + 48S** | **1KGP + 96S** | **1KGP + 384S** | **1KGP + 1285S** | **1KGP + 1285S in-house phasing** | **1285S** | **1285S in-house phasing** | **TOPMed-R3** |
| (0, 0.001) *127,857* | 3,528 3% | 11,893 9% | 17,681 14% | 26,389 21% | 48,321 38% | 72,363 57% | 72,440 57% | 76,003 59% | 66,630 52% | 17,609 14% |
| [0.001, 0.01) *54,076* | 2,862 5% | 9,884 18% | 15,293 28% | 22,080 41% | 36,446 67% | 45,716 85% | 45,917 85% | 45,179 84% | 42,836 79% | 10,474 19% |
| [0.01, 0.05) *24,740* | 3,386 14% | 12,333 50% | 17,460 71% | 21,294 86% | 24,211 98% | 24,578 99% | 24,579 99% | 24,286 98% | 24,420 99% | 13,460 54% |
| [0.05, 0.50) *36,023* | 12,467 35% | 29,348 81% | 33,544 93% | 35,298 98% | 35,933 99.9% | 35,983 99.9% | 35,984 99.9% | 35,950 99.9% | 35,930 99.9% | 29,008 81% |
| Total *242,696* | 22,243 9% | 63,458 26% | 83,978 35% | 105,061 43% | 144,911 60% | 178,640 74% | 178,920 74% | 181,418 75% | 169,816 70% | 70,551 29% |
| **CHR 21** | **Reference Panel** | | | | | | | | | |
| **MAF** ***# variants lost in 1KGP*** | **1KGP + 4S** | **1KGP + 24S** | **1KGP + 48S** | **1KGP + 96S** | **1KGP + 384S** | **1KGP + 1285S** | **1KGP + 1285S in-house phasing** | **1285S** | **1285S in-house phasing** | **TOPMed-R3** |
| (0, 0.001) *28,864* | 877 3% | 2,383 8% | 3,672 13% | 5,346 19% | 10,840 38% | 16,019 55% | 16,016 55% | 17,275 60% | 14,783 51% | 3,109 11% |
| [0.001, 0.01) *14,057* | 834 6% | 2,420 17% | 3,660 26% | 5,223 37% | 9,133 65% | 11,841 84% | 11,853 84% | 11,883 85% | 10,954 78% | 2,212 16% |
| [0.01, 0.05) *6,681* | 1,210 18% | 3,314 50% | 4,548 68% | 5,418 81% | 6,384 96% | 6,566 98% | 6,566 98% | 6,540 98% | 6,466 97% | 3,349 50% |
| [0.05, 0.50) *11,186* | 3,489 31% | 8,606 77% | 10,008 89% | 10,602 95% | 10,972 98% | 11,063 99% | 11,063 99% | 11,018 98% | 10,963 98% | 8,569 77% |
| Total *60,788* | 6,410 11% | 16,723 28% | 21,888 36% | 26,589 44% | 37,329 61% | 45,489 75% | 45,498 75% | 46,716 77% | 43,166 71% | 17,239 28% |

**Table S3. Consequence and variant groupings used to test enrichment of variants.**

| Sequence Ontology Term | Consequence Group | Impact Group |
|---|---|---|
| splice_acceptor_variant | Splice variant | HIGH |
| splice_donor_variant | Splice variant | HIGH |
| stop_gained | Coding variant | HIGH |
| frameshift_variant | Coding variant | HIGH |
| stop_lost | Coding variant | HIGH |
| start_lost | Coding variant | HIGH |
| transcript_amplification | Coding variant | HIGH |
| feature_elongation | Coding variant | HIGH |
| feature_truncation | Coding variant | HIGH |
| inframe_insertion | Coding variant | MODERATE |
| inframe_deletion | Coding variant | MODERATE |
| missense_variant | Coding variant | MODERATE |
| protein_altering_variant | Coding variant | MODERATE |
| splice_donor_5th_base_variant | Splice variant | LOW |
| splice_region_variant | Splice variant | LOW |
| splice_donor_region_variant | Splice variant | LOW |
| splice_polypyrimidine_tract_variant | Splice variant | LOW |
| incomplete_terminal_codon_variant | Coding variant | LOW |
| start_retained_variant | Coding variant | LOW |
| stop_retained_variant | Coding variant | LOW |
| synonymous_variant | Coding variant | LOW |
| coding_sequence_variant | Coding variant | MODIFIER |
| mature_miRNA_variant | Non-coding transcript variant | MODIFIER |
| 5_prime_UTR_variant | UTR | MODIFIER |
| 3_prime_UTR_variant | UTR | MODIFIER |
| non_coding_transcript_exon_variant | Non-coding transcript variant | MODIFIER |
| intron_variant | Intron variant | MODIFIER |
| non_coding_transcript_variant | Non-coding transcript variant | MODIFIER |
| coding_transcript_variant | Coding variant | MODIFIER |
| upstream_gene_variant | Up/Down-stream variant | MODIFIER |
| downstream_gene_variant | Up/Down-stream variant | MODIFIER |
| TF_binding_site_variant | Regulatory variant | MODIFIER |
| regulatory_region_ablation | Regulatory variant | MODIFIER |
| regulatory_region_amplification | Regulatory variant | MODIFIER |
| regulatory_region_variant | Regulatory variant | MODIFIER |
| intergenic_variant | Intergenic variant | MODIFIER |

**Table S4. Enrichment in variant consequence and impact for Samoan-specific imputation.** Enrichment of variants in each consequence and impact group that were uniquely imputed in a Samoan panel ($r^2 \geq 0.80$ in either 1285S or 1KGP+1285S and $r^2 <$ 0.80 or absent from both TOPMed-R3 and 1KGP) compared to shared variants ($r^2 \geq 0.80$ in either 1285S or 1KGP+1285S and either TOPMed-R3 or 1KGP) was calculated with a two-sided z test for the proportion of variants with the given annotation within the MAF stratum. Fold change is the proportion of variants in that group in the unique set divided by the proportion of variants in that group in the shared set. Bolded results indicate those with p-values < 0.0045 (i.e. 5% type 1 error with Bonferroni adjustment for 11 variant groups). These results are based on n=1,834 imputed individuals on chromosomes 5 and 21.

| **MAF bin** | **[0.001, 0.01)** | | **[0.01, 0.05)** | | **[0.05, 0.5)** | |
|---|---|---|---|---|---|---|
| | fold change | p-value | fold change | p-value | fold change | p-value |
| **Consequence** | | | | | | |
| Coding variant | 1.672 | **2.00E-22** | 1.189 | 1.91E-02 | 1.668 | **2.20E-07** |
| Splice variant | 1.289 | **1.92E-03** | 1.200 | 1.23E-01 | 1.403 | 3.14E-02 |
| UTR | 1.216 | **9.82E-11** | 1.056 | 2.25E-01 | 1.246 | **3.64E-04** |
| Up/Down-stream variant | 1.084 | **1.39E-07** | 1.006 | 7.96E-01 | 1.342 | **3.97E-26** |
| Intron variant | 0.975 | **8.42E-16** | 0.986 | **2.16E-03** | 0.934 | **6.59E-23** |
| Non-coding transcript variant | 1.102 | **3.90E-06** | 1.285 | **1.34E-15** | 1.242 | **2.21E-07** |
| Intergenic variant | 0.996 | 5.84E-01 | 0.985 | 2.05E-01 | 1.005 | 7.52E-01 |
| **Impact** | 1.484 | 6.43E-02 | 2.065 | 2.31E-02 | 2.773 | **1.33E-03** |
| HIGH | 1.780 | **2.91E-16** | 1.214 | 4.31E-02 | 1.461 | 1.07E-02 |
| MODERATE | 1.410 | **7.09E-09** | 1.133 | 1.45E-01 | 1.576 | **1.98E-05** |
| LOW | 0.996 | **3.07E-23** | 0.998 | 4.97E-03 | 0.996 | **3.80E-08** |
| MODIFIER | 1.672 | **2.00E-22** | 1.189 | 1.91E-02 | 1.668 | **2.20E-07** |

**Table S5. Enrichment in variant consequence and impact for Samoan-specific imputation.** Enrichment of variants in each consequence and impact group that were uniquely imputed in a Samoan panel ($r^2 \geq 0.80$ in either 1285S or 1KGP+1285S and $r^2 <$ 0.80 or absent from both TOPMed-R3 and 1KGP) compared to shared variants ($r^2 \geq 0.80$ in either 1285S or 1KGP+1285S and either TOPMed-R3 or 1KGP) was calculated with a two-sided z test for the proportion of variants with the given annotation within the MAF stratum. Fold change is the proportion of variants in that group in the unique set divided by the proportion of variants in that group in the shared set. Bolded results indicate those with p-values < 0.0045 (i.e. 5% type 1 error with Bonferroni adjustment for 11 variant groups). These results are based on n=100 imputed individuals across all autosomes. P-values listed as 0 were less than 2.225074e-308.

| **MAF bin** | **[0.001, 0.01)** | | **[0.01, 0.05)** | | **[0.05, 0.5)** | |
|---|---|---|---|---|---|---|
| | fold change | p-value | fold change | p-value | fold change | p-value |
| **Consequence** | | | | | | |
| Coding variant | 1.708 | **4.32E-152** | 1.635 | **1.15E-162** | 1.745 | **0.00E+00** |
| Splice variant | 1.315 | **1.43E-07** | 1.309 | **2.02E-09** | 1.484 | **8.72E-38** |
| UTR | 1.304 | **3.06E-68** | 1.276 | **1.56E-77** | 1.394 | **3.69E-277** |
| Up/Down-stream variant | 1.107 | **4.89E-62** | 1.103 | **4.29E-79** | 1.300 | **0.00E+00** |
| Intron variant | 1.010 | **3.24E-08** | 1.001 | 4.59E-01 | 0.982 | **2.00E-54** |
| Non-coding transcript variant | 1.134 | **3.38E-20** | 1.143 | **2.04E-29** | 1.299 | **9.18E-211** |
| Intergenic variant | 0.937 | **3.95E-170** | 0.951 | **7.38E-135** | 0.925 | **0.00E+00** |
| **Impact** | | | | | | |
| HIGH | 1.957 | **3.74E-08** | 1.435 | **3.55E-04** | 1.726 | **1.59E-14** |
| MODERATE | 2.038 | **1.29E-144** | 1.829 | **9.76E-133** | 1.897 | **1.17E-293** |
| LOW | 1.324 | **9.91E-26** | 1.392 | **2.08E-44** | 1.562 | **4.25E-170** |
| MODIFIER | 0.995 | **1.51E-154** | 0.996 | **9.45E-167** | 0.995 | **0.00E+00** |

**Table S6. Mean $r^2$ by reference panel.** Mean $r^2$ is given by minor allele frequency (MAF) based on the protocol 8 imputation across reference panels and chromosome.

| CHR 5 | Reference Panel | | | | | | | | | |
|---|---|---|---|---|---|---|---|---|---|---|
| MAF | 1KGP | 1KGP + 4S | 1KGP + 24S | 1KGP + 48S | 1KGP + 96S | 1KGP + 384S | 1KGP + 1285S | 1KGP + 1285S in-house phasing | 1285S | 1285S in-house phasing | TOPMed-R3 |
| (0, 0.001) | 0.538 | 0.551 | 0.585 | 0.609 | 0.641 | 0.712 | 0.778 | 0.779 | 0.779 | 0.726 | 0.582 |
| [0.001, 0.01) | 0.710 | 0.727 | 0.772 | 0.804 | 0.840 | 0.907 | 0.947 | 0.948 | 0.940 | 0.916 | 0.726 |
| [0.01, 0.05) | 0.791 | 0.817 | 0.888 | 0.926 | 0.954 | 0.979 | 0.988 | 0.989 | 0.985 | 0.983 | 0.886 |
| [0.05, 0.1) | 0.859 | 0.894 | 0.952 | 0.968 | 0.978 | 0.988 | 0.993 | 0.993 | 0.991 | 0.991 | 0.938 |
| [0.1, 0.2) | 0.910 | 0.936 | 0.970 | 0.979 | 0.985 | 0.992 | 0.995 | 0.996 | 0.994 | 0.994 | 0.956 |
| [0.2, 0.3) | 0.937 | 0.954 | 0.976 | 0.983 | 0.988 | 0.993 | 0.996 | 0.996 | 0.995 | 0.995 | 0.961 |
| [0.3, 0.4) | 0.947 | 0.960 | 0.978 | 0.985 | 0.989 | 0.994 | 0.997 | 0.997 | 0.996 | 0.995 | 0.963 |
| [0.4, 0.5) | 0.950 | 0.963 | 0.980 | 0.986 | 0.990 | 0.995 | 0.997 | 0.997 | 0.996 | 0.996 | 0.966 |

| CHR 21 | Reference Panel | | | | | | | | | |
|---|---|---|---|---|---|---|---|---|---|---|
| MAF | 1KGP | 1KGP + 4S | 1KGP + 24S | 1KGP + 48S | 1KGP + 96S | 1KGP + 384S | 1KGP + 1285S | 1KGP + 1285S in-house phasing | 1285S | 1285S in-house phasing | TOPMed-R3 |
| (0, 0.001) | 0.466 | 0.482 | 0.515 | 0.542 | 0.576 | 0.668 | 0.740 | 0.740 | 0.748 | 0.692 | 0.522 |
| [0.001, 0.01) | 0.645 | 0.666 | 0.717 | 0.753 | 0.792 | 0.882 | 0.934 | 0.934 | 0.932 | 0.892 | 0.678 |
| [0.01, 0.05) | 0.736 | 0.782 | 0.863 | 0.908 | 0.937 | 0.97 | 0.982 | 0.983 | 0.98 | 0.976 | 0.859 |
| [0.05, 0.1) | 0.819 | 0.864 | 0.932 | 0.954 | 0.967 | 0.982 | 0.990 | 0.990 | 0.988 | 0.985 | 0.915 |
| [0.1, 0.2) | 0.886 | 0.918 | 0.959 | 0.971 | 0.978 | 0.988 | 0.993 | 0.993 | 0.992 | 0.990 | 0.946 |
| [0.2, 0.3) | 0.909 | 0.933 | 0.965 | 0.974 | 0.981 | 0.99 | 0.994 | 0.994 | 0.993 | 0.992 | 0.951 |
| [0.3, 0.4) | 0.922 | 0.941 | 0.969 | 0.977 | 0.983 | 0.991 | 0.995 | 0.995 | 0.994 | 0.993 | 0.956 |
| [0.4, 0.5) | 0.930 | 0.947 | 0.972 | 0.979 | 0.985 | 0.991 | 0.995 | 0.995 | 0.994 | 0.993 | 0.959 |

**Table S7. Median $r^2$ by reference panel.** Median $r^2$ is given by minor allele frequency (MAF) based on the protocol #8 imputation across reference panels and chromosome.

| CHR 5 | Reference Panel | | | | | | | | | |
|---|---|---|---|---|---|---|---|---|---|---|
| MAF | 1KGP | 1KGP + 4S | 1KGP + 24S | 1KGP + 48S | 1KGP + 96S | 1KGP + 384S | 1KGP + 1285S | 1KGP + 1285S in-house phasing | 1285S | 1285S in-house phasing | TOPMed-R3 |
| (0, 0.001) | 0.536 | 0.576 | 0.675 | 0.739 | 0.810 | 0.920 | 0.982 | 0.982 | 0.979 | 0.944 | 0.622 |
| [0.001, 0.01) | 0.879 | 0.896 | 0.933 | 0.950 | 0.964 | 0.986 | 0.995 | 0.995 | 0.992 | 0.987 | 0.797 |
| [0.01, 0.05) | 0.932 | 0.949 | 0.974 | 0.983 | 0.988 | 0.994 | 0.997 | 0.997 | 0.996 | 0.996 | 0.944 |
| [0.05, 0.1) | 0.963 | 0.971 | 0.986 | 0.990 | 0.993 | 0.997 | 0.998 | 0.998 | 0.998 | 0.998 | 0.968 |
| [0.1, 0.2) | 0.975 | 0.981 | 0.990 | 0.993 | 0.995 | 0.998 | 0.999 | 0.999 | 0.998 | 0.998 | 0.973 |
| [0.2, 0.3) | 0.978 | 0.984 | 0.991 | 0.994 | 0.996 | 0.998 | 0.999 | 0.999 | 0.998 | 0.998 | 0.975 |
| [0.3, 0.4) | 0.981 | 0.985 | 0.992 | 0.994 | 0.996 | 0.998 | 0.999 | 0.999 | 0.998 | 0.998 | 0.976 |
| [0.4, 0.5) | 0.980 | 0.985 | 0.992 | 0.994 | 0.996 | 0.998 | 0.999 | 0.999 | 0.998 | 0.998 | 0.976 |
| **CHR 21** | **Reference Panel** | | | | | | | | | | |
| **MAF** | **1KGP** | **1KGP + 4S** | **1KGP + 24S** | **1KGP + 48S** | **1KGP + 96S** | **1KGP + 384S** | **1KGP + 1285S** | **1KGP + 1285S in-house phasing** | **1285S** | **1285S in-house phasing** | **TOPMed-R3** |
| (0, 0.001) | 0.369 | 0.404 | 0.497 | 0.574 | 0.669 | 0.864 | 0.961 | 0.961 | 0.965 | 0.919 | 0.509 |
| [0.001, 0.01) | 0.755 | 0.795 | 0.863 | 0.901 | 0.929 | 0.973 | 0.990 | 0.990 | 0.990 | 0.978 | 0.725 |
| [0.01, 0.05) | 0.871 | 0.928 | 0.960 | 0.973 | 0.980 | 0.992 | 0.996 | 0.996 | 0.995 | 0.995 | 0.923 |
| [0.05, 0.1) | 0.922 | 0.947 | 0.973 | 0.981 | 0.987 | 0.994 | 0.997 | 0.997 | 0.996 | 0.996 | 0.949 |
| [0.1, 0.2) | 0.960 | 0.971 | 0.985 | 0.990 | 0.993 | 0.996 | 0.998 | 0.998 | 0.998 | 0.997 | 0.967 |
| [0.2, 0.3) | 0.964 | 0.972 | 0.985 | 0.990 | 0.993 | 0.996 | 0.998 | 0.998 | 0.997 | 0.996 | 0.967 |
| [0.3, 0.4) | 0.969 | 0.977 | 0.987 | 0.991 | 0.994 | 0.997 | 0.998 | 0.998 | 0.997 | 0.997 | 0.970 |
| [0.4, 0.5) | 0.972 | 0.980 | 0.989 | 0.992 | 0.994 | 0.997 | 0.998 | 0.998 | 0.997 | 0.997 | 0.971 |

**Table S8. Median $F_{ST}$ and LD score by minor allele frequency (MAF) and chromosome.** Weir and Cockerham $F_{ST}$ and LD score were calculated on the whole-genome sequencing data comprising the 1KGP + 1285S reference panel. Both $F_{ST}$ and LD score were stratified into 'low' and 'high' about the median within each MAF bin.

| MAF | CHR 5 | | CHR 21 | |
|---|---|---|---|---|
| | median $F_{ST}$ | median LD score | median $F_{ST}$ | median LD score |
| (0, 0.001) | 0.022 | 56.1 | 0.022 | 44.7 |
| [0.001, 0.01) | 0.063 | 125.3 | 0.065 | 105.8 |
| [0.01, 0.05) | 0.049 | 124.7 | 0.050 | 114.3 |
| [0.05, 0.1) | 0.054 | 137.0 | 0.055 | 117.3 |
| [0.1, 0.2) | 0.067 | 158.3 | 0.067 | 144.4 |
| [0.2, 0.3) | 0.072 | 182.4 | 0.075 | 158.6 |
| [0.3, 0.4) | 0.083 | 203.3 | 0.076 | 162.6 |
| [0.4, 0.5) | 0.086 | 206.5 | 0.083 | 181.5 |

**Table S9. Number of well-imputed variants by reference panel for meta imputation.** Number of well-imputed variants ($r^2 \geq 0.80$) by minor allele frequency (MAF) and chromosome for TOPMed-R3, 1285S and their meta imputation.

| **CHR 5** | | | | | |
|---|---|---|---|---|---|
| | | | **MAF bin** | | |
| **Reference Panel** | **(0, 0.001)** | **[0.001, 0.01)** | **[0.01, 0.05)** | **[0.05, 0.50)** | **Total** |
| TOPMed-R3 | 155,605 | 82,367 | 82,544 | 338,837 | 659,353 |
| 1285S | 218,877 | 169,993 | 106,796 | 365,428 | 861,094 |
| Meta imputation | 261,237 | 159,502 | 105,074 | 365,194 | 891,007 |
| **CHR 21** | | | | | |
| | | | **MAF bin** | | |
| **Reference Panel** | **(0, 0.001)** | **[0.001, 0.01)** | **[0.01, 0.05)** | **[0.05, 0.50)** | **Total** |
| TOPMed-R3 | 25,364 | 15,436 | 16,752 | 73,091 | 130,643 |
| 1285S | 41,437 | 35,764 | 22,934 | 79,469 | 179,604 |
| Meta imputation | 49,273 | 33,772 | 22,617 | 79,784 | 185,446 |
